# Fetal pathogenesis of scoliosis suggested by asymmetry of gene expression in paravertebral muscles

**DOI:** 10.1101/2025.11.11.25339986

**Authors:** Tian Cheng, Gamze Yazgeldi Gunaydin, Elisabet Einarsdottir, Sini Ezer, Juha Kere, Shintaro Katayama, Paul Gerdhem

## Abstract

**Background:** The malfunction of paravertebral muscles may contribute to the development of idiopathic scoliosis. Several candidate genes have been linked to scoliosis, but the underlying mechanisms and cell types remain unclear.

**Methods:** We included 40 idiopathic scoliosis cases and 19 controls. Muscle biopsies were obtained bilaterally in the cases, and at least unilaterally in the controls. RNA sequencing, differential gene expression analysis and gene set enrichment analysis were performed between cases and controls, and between convex and concave sides in the cases. Hounsfield units (HU) in paravertebral muscles from preoperative CT were assessed at biopsy sites in cases.

**Results:** Qualified transcriptome analysis included 56 samples (30 convex, 26 concave) from 35 scoliosis cases and 22 samples from 17 controls. Among 14,212 expressed genes included in the downstream analysis, 22 differentially expressed genes were identified between cases and controls and 16 between convex and concave sides. Scoliosis cases showed decreased fetal muscle and immune cells. Convex side showed decreased fibro-adipogenic progenitors, endothelial cell types, satellite cells and myeloid cells and increased fetal skeletal muscle cells. Morphological asymmetry was verified by the HU findings.

**Conclusion:** These results indicated asymmetry of gene expression profiles and suggests a fetal developmental origin of idiopathic scoliosis, with increased muscle mass pushing convexity.

## Introduction

Idiopathic scoliosis is the most prevalent type of spinal deformity, accounting for 80% of all scoliosis cases. Its prevalence is approximately 2-3%, and it is more common in females than in males (1, 2). The progression can be rapid during a child’s growth spurt, and if left untreated, a large curve can lead to respiratory dysfunction and ultimately premature death (3). Brace treatment or surgery with spinal correction and fusion is indicated to halt progression (4, 5).

Idiopathic scoliosis is considered a polygenic and multifactorial disease, but its etiology and pathogenesis remain poorly understood (5). Twin and family studies suggest a strong genetic component (6–8), although currently known single nucleotide variants explain only a small fraction of the underlying genetics (9).

Several theories have been proposed regarding the mechanism underlying the development and progression of idiopathic scoliosis (5). The inherent shape of the spine may be part of the susceptibility of scoliosis (10). The paravertebral muscles play a crucial role in spinal stability, and imbalance in these muscles may contribute to the development of idiopathic scoliosis (5). The convex side has larger muscle volume and type I muscle fibers, while the concave side exhibits greater fatty infiltration and type II fibers, according to both histological and radiological studies (11).

These observations led us to hypothesize that there might be differences in gene expression related to muscle and adipose tissue development between the concave and convex sides of the scoliotic spine, as well as between scoliosis cases and controls without scoliosis. We therefore aimed to study gene expression differences, with special emphasis on pathways related to muscle, adipose, cartilage and bone development.

## Results

### Patient characteristics

35 scoliosis cases (24 females) and 17 controls (5 females) were included (Figure 1) in the final analysis. Mean age (SD) at surgery was 15.9 (2.4) years for scoliosis cases and 17.1 (2.2) years for controls. The mean major curve size Cobb angle in the scoliosis cases was 59 (10) degrees, and major curve types were thoracic (n=25), thoracolumbar (n=5), and lumbar (n=5). Sampling level ranged from the T5/T6 disc level to the L4 vertebra for the scoliosis cases and from the T11/T12 disc level to the S1 vertebra in the controls (Supplementary Table 1).

**Figure 1.**
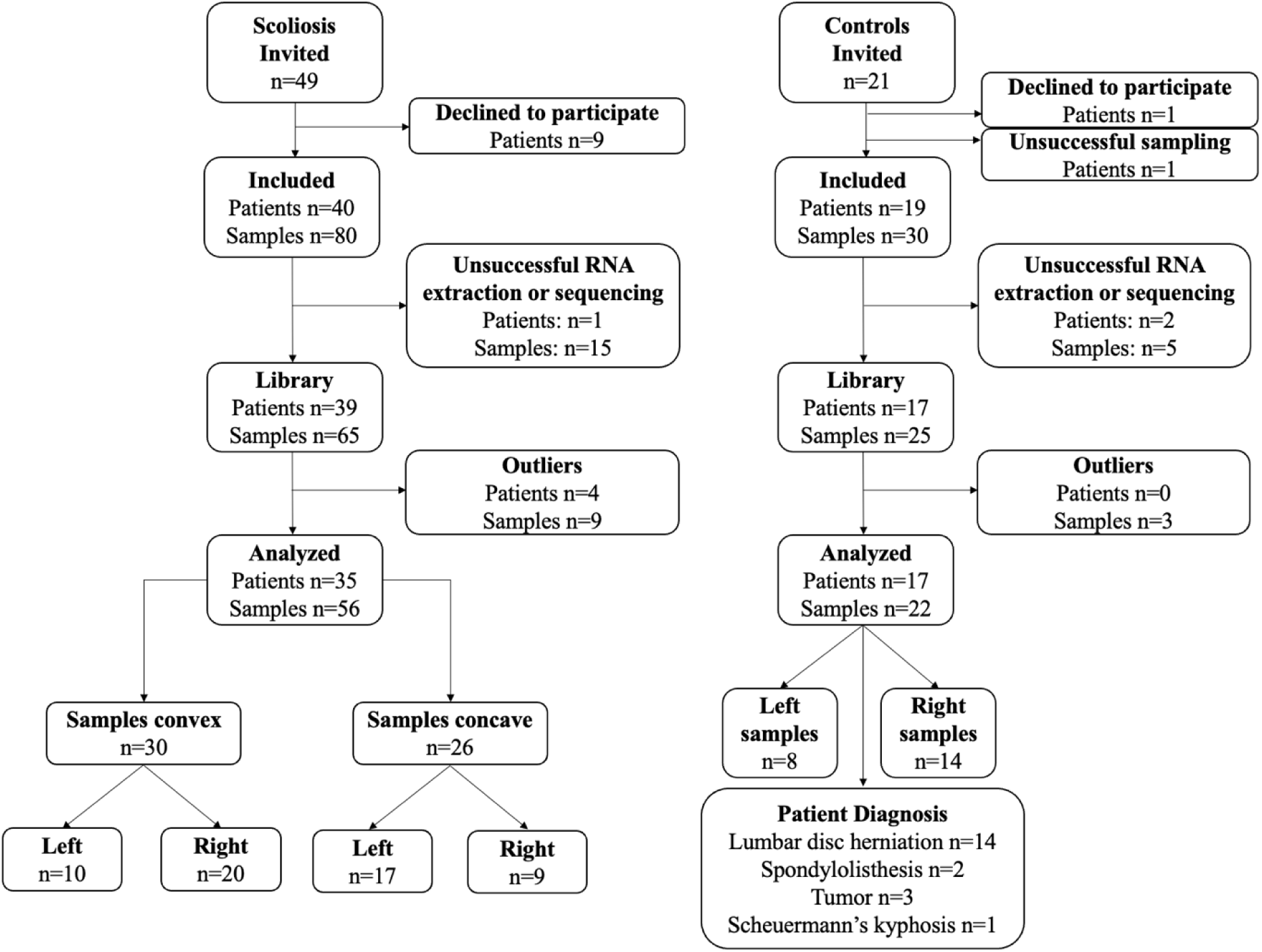
Flowchart of patient inclusion and exclusion.

### Radiological analysis

Mean Hounsfield unit (HU) of the paravertebral muscle at the level of sampling on the convex side and concave side is presented in Figure 2. The convex side HU was significantly higher, 49 (8) compared to the concave side, 41 (10), p=0.0037 suggesting more muscle tissues on the convex side and more adipose tissue on the concave side.

**Figure 2:**
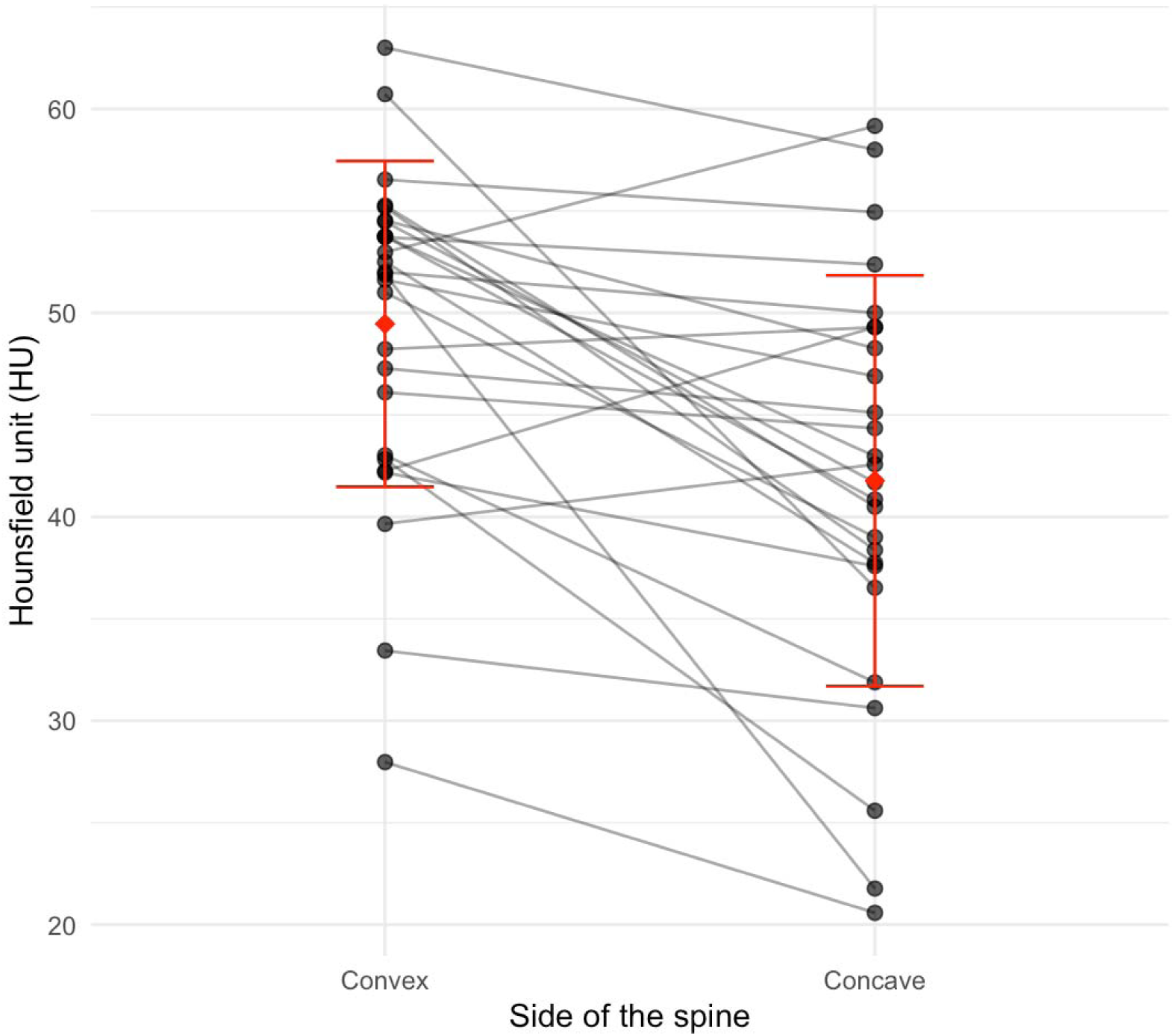
Paired dot plot showing Hounsfield unit (HU) of the parvertebral muscle at the level of sampling.

### Overview of gene expression profiling

After removing outliers during quality control, 78 of the initial 96 samples were kept for further analysis (Supplementary Figure 1). Of these, 56 muscle samples (26 concave and 30 convex) were from the scoliosis cases group, and 22 samples were from the control group.

After removing genes with low expression levels, a total of 14,212 genes were retained for downstream analysis. Among these, 7,807 fluctuating genes were identified based on their higher variability compared to technical variation of added spike-in synthetic RNAs, and their adjusted p-values (padj) were used for further analyses. Principal component analysis (PCA) showed a bias between the two STRT libraries, which was still present after library bias correction (Supplementary Figure 2). Therefore, library information was included as a covariate factor in the differential gene expression (DGE) model.

### Scoliosis muscle shows shift in immune and progenitor cell signature

In the comparison between scoliosis cases and control groups, 22 significant differentially expressed genes (DEGs) were identified (padj < 0.05 and fluctuation padj < 0.05, (Figure 3a), including genes associated with extracellular matrix remodeling *(ANGPTL7, COMP, RASD1, LUM, PTN, PLAU, INHBA),* immune regulation and stress responses *(JUNB, IDO1, C2, S100A13, NQO1, LGMN, NNMT),* developmental regulation *(INHBA, SYCE1, SLFN5),* and cell proliferation *(CNN1, PTN, TF, JPT1, SLFN5).* In addition, fetal hematopoiesis *(HBG1)* was represented (Supplementary Table 2).

**Figure 3.**
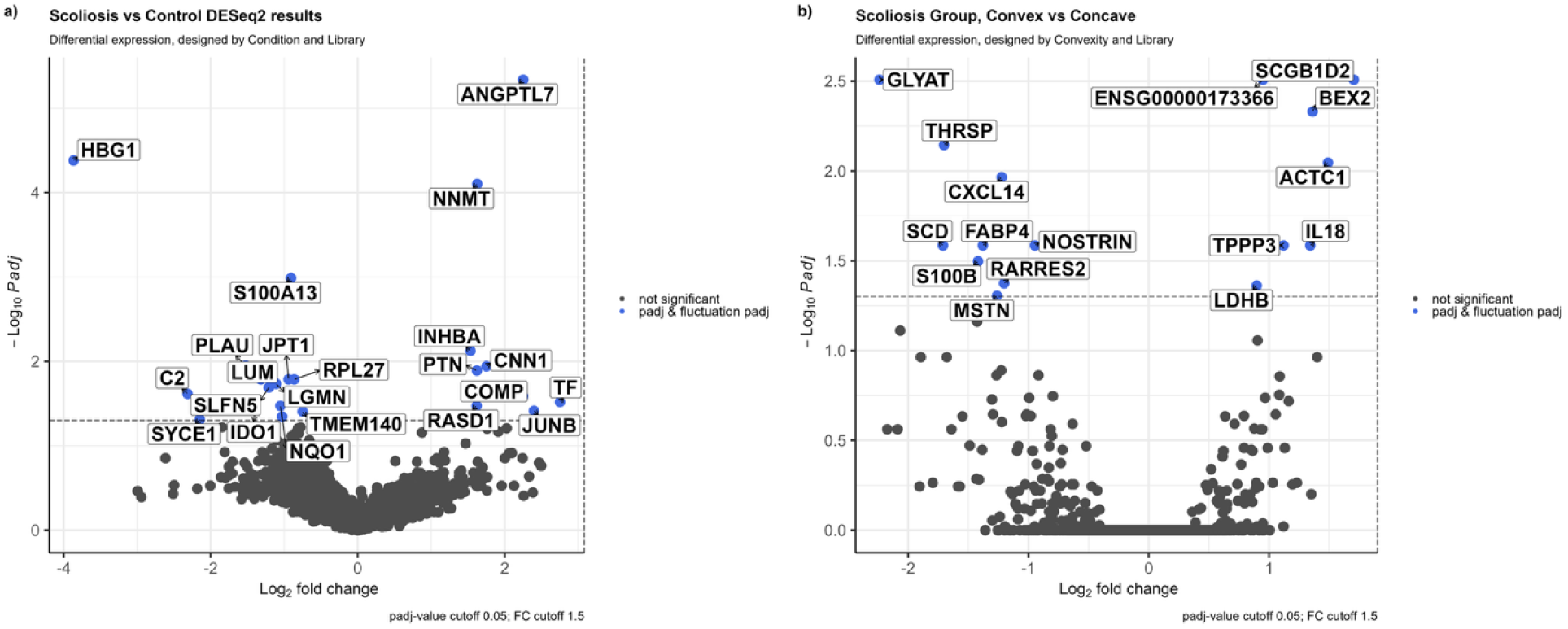
Gene expression profiles in scoliosis vs. control and convex vs. concave conditions. (A) Comparison between scoliosis and control samples, with genes highlighted in blue if they have both padj<0.05 and fluctuation padj<0.05, including significantly changed and fluctuated genes. (B) Comparison between convex and concave conditions, with significantly changed genes (padj<0.05 and fluctuation padj<0.05) also highlighted in blue. The horizontal dashed line at −log10(0.05)=1.301 shows the significance threshold for the padj. Gray dots are genes that ar not statistically significant with padj>0.05 and |log2FC|<1.5.

To test whether the transcriptomic changes in scoliosis are linked to specific cell types, we performed cell type enrichment analysis using DEGs. The gene set enrichment analysis (GSEA) showed a general decrease in immune and progenitor cell signatures in non-fetal scoliosis muscle, while fetal Schwann cell signatures were increased (Table 1). These results indicate that scoliosis muscle is characterized by changes in immune processes and altered cell-population signatures.

**Table 1.** Enriched cell types in scoliosis versus control comparison. This table shows cell type ontology analysis results of all genes, as well as upregulated (log2FC>0) in scoliosis and downregulated (log2FC<0) based on log2FC and p-value.

| Direction | Cell types | padj | NES |
| --- | --- | --- | --- |
| Down in scoliosis |  |  |  |
|  | Skeletal muscle T cells | 2.53e-10 | -1.68 |
|  | Skeletal muscle B cells | 2.20e-07 | -1.58 |
|  | Skeletal muscle satellite cells | 2.87e-05 | -1.38 |
|  | Skeletal muscle FBN1 FAP cells | 4.26e-04 | -1.36 |
|  | Skeletal muscle myeloid cells | 6.00e-04 | -1.31 |
|  | Skeletal muscle PCV endothelial cells | 1.77e-03 | -1.35 |
|  | Skeletal muscle NK cells | 0.012 | -1.30 |
|  | Fetal muscle skeletal muscle cells | 0.013 | -1.33 |
|  | Skeletal muscle FAP cells | 0.036 | -1.28 |
| Up in scoliosis |  |  |  |
|  | Fetal muscle Schwann cells | 3.77e-06 | 2.22 |

padj =Adjusted p-value using Benjamini–Hochberg correction for multiple hypothesis testing, NES = Normalized Enrichment Score, FAP = Fibro-Adipogenic Progenitors, PCV= Post-Capillary Venule.

### Convex-concave asymmetry reflects developmental imbalance

In the convex versus concave comparison, 16 DEGs were identified (padj < 0.05 and fluctuation padj < 0.05) (Figure 3b), including genes that regulate muscle cell structure and regeneration *(ACTC1, MSTN, S100B, TPP3)*, adipose remodeling *(FABP4, THRSP, RARRES2, SCD)*, inflammatory mediators *(CXCL14, IL18, NOSTRIN),* metabolic genes *(GLYAT, LDHB)*, and cell cycle/apoptosis regulators *(BEX2)* (Supplementary Table 2). On the convex side, gene signatures related to adult skeletal muscle cell types (fibro-adipogenic progenitors (FAP), Post-Capillary Venule (PCV) endothelial cells, endothelial cells, FBN1 FAP cells, and satellite cells from Rubenstein dataset) were markedly downregulated, whereas fetal skeletal muscle cell types (vascular endothelial cells, skeletal muscle cells, and myeloid cells from DESCARTES dataset) were significantly enriched (Table 2). Among fetal muscle related signatures, vascular endothelial cells and myeloid cells were downregulated, while skeletal muscle cells were upregulated. This suggests that scoliosis asymmetry may involve a selective activation of muscle-related developmental programs on the convex in contrast to the concave side, which may contribute to structural imbalance in scoliosis.

**Table 2.** Enriched cell types in convex versus concave comparison. This table shows cell type ontology analysis results of all genes, as well as upregulated (log2FC>0) in convex and downregulated (log2FC<0) based on log2FC and p-value.

| Direction | Cell types | padj | NES |
| --- | --- | --- | --- |
| Down in convex |  |  |  |
|  | Skeletal muscle FAP cells | 9.45e-08 | -2.02 |
|  | Skeletal muscle PCV endothelial cells | 4.94e-05 | -1.759 |
|  | Fetal muscle vascular endothelial cells | 5.38e-05 | -2.06 |
|  | Skeletal muscle endothelial cells | 3.04e-04 | -1.75 |
|  | Skeletal muscle FBN1 FAP cells | 3.12e-03 | -1.54 |
|  | Skeletal muscle satellite cells | 0.007 | -1.45 |
|  | Fetal muscle myeloid cells | 0.028 | -1.46 |
| Up in convex |  |  |  |
|  | Fetal muscle skeletal muscle cells | 2.31e-03 | 1.62 |

padj = Adjusted p-value using Benjamini–Hochberg correction for multiple hypothesis testing, NES = Normalized Enrichment Score, FAP = Fibro-Adipogenic Progenitors, PCV= Post-Capillary Venule.

### Differentially expressed genes show developmental stage specific and cell population specific expression patterns

Cell-type enrichment analysis suggested that muscle-resident progenitor cells were enriched in both cell-type enrichment analyses (Table 1 and 2). To explore whether this enrichment is associated with developmental regulation, we next examined expression of differently changed genes across human developmental stages (12, 13). A heatmap showing expression values across embryonic, fetal, juvenile, and adult muscle tissues showed that the majority of DEGs show dynamic expression patterns during early development (Supplementary Figure 3). Notably, *RPL27, LDHB, PTN, HN1, LUM,* and *SCD* showed high expression in embryonic and fetal stages, whereas *CXCL14, JUNB, NNMT,* and *RASD1* had higher expression in later stages.

To further explore the cellular specificity of these patterns, we generated violin plots separated by both developmental stage and cell type (Figure 4). Among the DEGs visualized, *PTN, LUM, RPL27, JUNB, CXCL14, LDHB,* and *S100A13* showed notable expression in mesenchymal stem cells (MSC) during fetal development. Additionally, *RPL27, JUNB, CXCL14, S100A13,* and *NNMT* showed strong expression in FAP. Genes such as *FABP4, NNMT, INHBA, RASD1, BEX2, SCD, NOSTRIN,* and *RARRES2* showed more cell population specific expression patterns across endothelial cells, hematopoietic cells, and Schwann cell populations. Genes such as *LGMN, PLAU, NQO1, CAT,* and *MSTN* showed very low expression (Figure 4).

**Figure 4.**
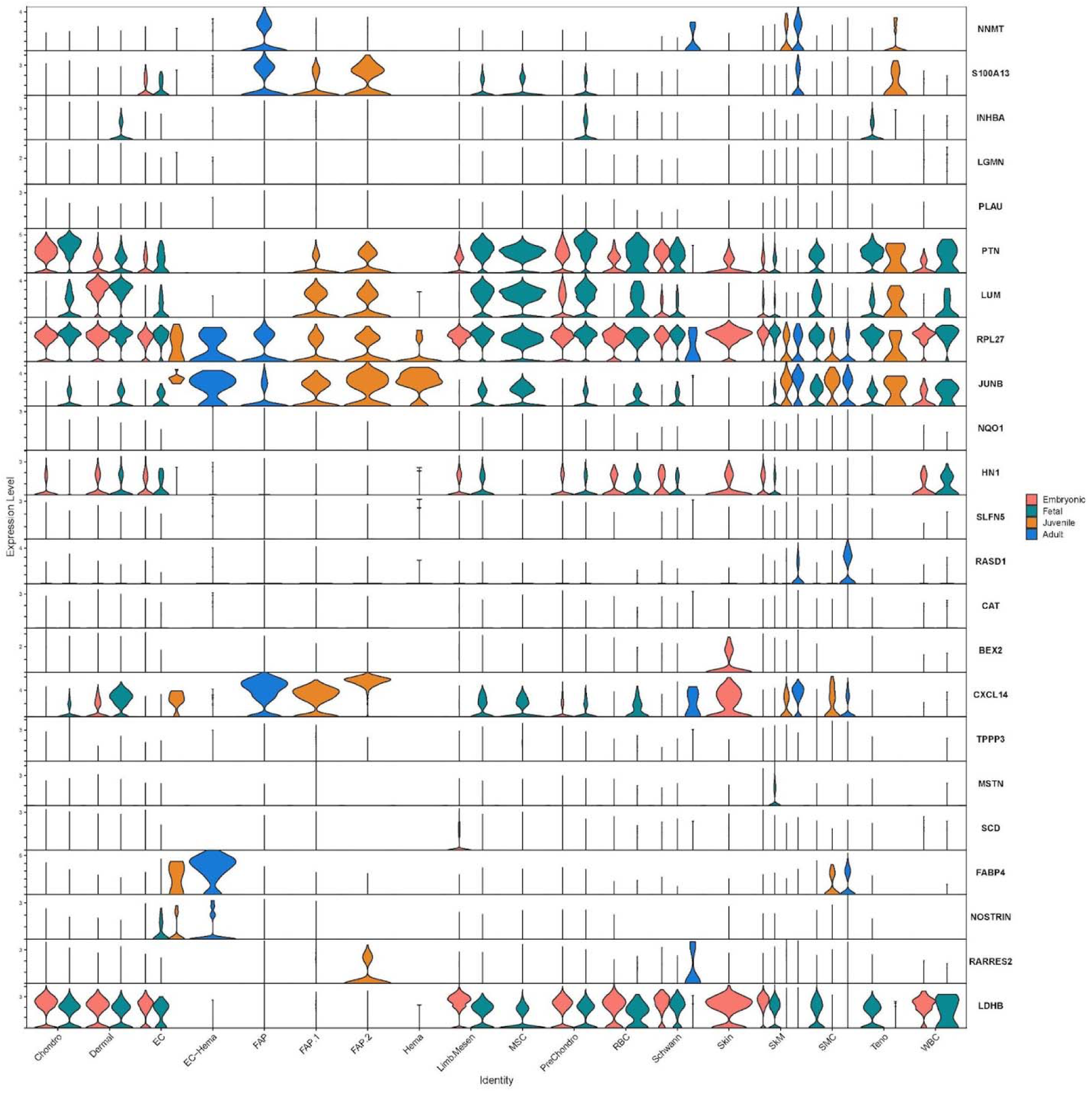
Expression of DEGs in UCSC-Cell Browser Skeletal Muscle Dataset. Stacked violin plot showing log1p normalized expression of our DEGs across different cell types and developmental stages. Cell types are Chondro (Chondrocytes), PreChondro (Pre-Chondrocytes), Limb.Mesen (Limb Mesenchymal Cells), MSC (Mesenchymal Stem Cells), FAP (Fibro-Adipogenic Progenitors), Hema (Hematopoietic Cells), RBC (Red Blood Cells), WBC (White Blood Cells), Skin, Dermal, SkM (Skeletal Muscle Cells), SMC (Smooth Muscle Cells), Schwann, EC (Endothelial), and Teno (Tenocytes). Developmental stages are colored a embryonic (pink), fetal (green), juvenile (orange), and adult (blue).

These observations suggest that scoliosis-associated transcriptional changes involve both side-specific and developmental regulation. Such regulation is potentially driven by enrichment of several DEGs in fetal mesenchymal and progenitor cell populations such as FAPs, as well as differentiating or satellite muscle cells.

## Discussion

Our study showed clear differences in gene regulation between the convex and concave sides. While the convex side was enriched for fetal skeletal muscle signatures, the concave side was enriched for adipogenic signatures (Table 2). Although these asymmetrical composition patterns between the two paravertebral muscle sides have been reported previously (11), our data extend these observations by suggesting mechanistic insight into their developmental timing and transcriptional regulation.

The gene expression changes behind this asymmetry mainly involved fat-related genes linked to fat accumulation on the concave side and muscle-related genes linked to muscle activity on the convex side. *FABP4*, a key adipocyte marker, was upregulated on the concave side in our dataset, consistent with findings from Shu *et al.* (14), which showed increased *FABP4* in the paravertebral muscles of idiopathic scoliosis cases, along with impaired muscle regeneration. Similarly, *THRSP*, a regulator of *de novo* lipogenesis, was also upregulated on the concave side in our data. This is consistent with findings from Wang *et al.* (15), which showed increased *THRSP* expression during adipogenic differentiation of mesenchymal stem cells in adolescent idiopathic scoliosis cases. *RARRES2*, another adipokine, was upregulated on the concave side in our data. Although its direct role in scoliosis is not clear, it is known to promote adipocyte differentiation via CMKLR1 signaling (16) and it impairs insulin signaling in skeletal muscle, potentially affecting muscle function (17). Moreover, increased circulating chemerin levels have also been observed in lipodystrophy, a condition characterized by adipose tissue remodeling (18). *ACTC1*, a structural muscle gene, was upregulated on the convex side, and *MSTN*, a negative regulator of muscle growth, was downregulated on the concave side, suggesting different muscle activation patterns between the two sides. These transcriptomic findings are consistent with previous radiological observations from Jiang *et al.* who found that paravertebral muscles have greater volume on the convex side and increased fatty infiltration on the concave side in adolescent idiopathic scoliosis cases (11). The normal paravertebral muscle Hounsfield Unit (HU) on the CT ranges from 40-50. In muscles with fatty infiltration, the HU is decreased since adipose tissue has a much lower HU than muscle tissues. In our radiological analysis, the HU measurement was significantly lower on the concave side compared to the convex side of the scoliosis spine, supporting the theory of more fatty infiltration on the concave side.

We also observed gene expression changes linked to extracellular matrix composition and tissue remodeling. For example, *COMP*, an extracellular cartilage oligomeric matrix protein, was upregulated in paravertebral muscle of scoliosis cases, whereas previous studies have found lower gene expression or serum levels in scoliosis (19–21). Also, *LUM,* an extracellular matrix proteoglycan and key regulator of collagen fibrillogenesis, was downregulated in scoliosis cases compared to controls. This is consistent with previous findings showing that disrupted *LUM* function is linked to impaired collagen organization and extracellular matrix disorganization in scoliotic discs (22). *ANGPTL7*, a regulator of extracellular matrix organization, was significantly upregulated in the scoliosis cases compared to controls in our dataset. Although its role in scoliosis has not been directly studied, findings from other skeletal conditions suggest potential relevance. Liu *et al.* (23) showed that ANGPTL7+ chondrocytes promote angiogenesis via FGF2-FGFR2 signaling and contribute to cartilage degeneration in a mouse model of knee osteoarthritis. Together, these findings highlight dysregulation of extracellular matrix remodeling in scoliosis, in concordance with recent reports (24) (REF).

The findings raise the question of whether transcriptional changes are the result of the scoliosis itself or vice versa. One theory suggests the involvement of progenitor cells in embryogenesis and fetal development, resulting in an imbalance between adipocytes, myoblasts, chondrocytes and osteoblasts and eventually to the phenotype scoliosis (5, 25). To identify specific cell populations contributing to the observed gene expression patterns, we performed cell population specific enrichment analysis using fetal (26) and adult muscle datasets (27).

In the scoliosis versus control comparison, we observed downregulation of several immune cell types including T cells, B cells, satellite cells, FBN1 FAP cells, myeloid cells, PCV endothelial cells, NK cells, FAPs (from the adult skeletal muscle dataset by Rubenstein *et al.*). In contrast, fetal Schwann cells were upregulated, and fetal skeletal muscle cells were downregulated (from the fetal dataset by Cao *et al.*) (Table 1).

In the convex versus concave comparison, we observed a similar trend with downregulation of FAPs, PCV endothelial cells, endothelial cells, FBN1 FAP cells, and satellite cells (from the adult skeletal muscle dataset by Rubenstein *et al.*). In contrast, fetal vascular endothelial cells and fetal myeloid cells were downregulated, and fetal skeletal muscle cells were upregulated (from the fetal dataset by Cao *et al.*) (Table 2). Supporting this, Dai *et al.* (28) found that a shift toward adipogenic differentiation in congenital scoliosis may be triggered by epigenetic modulation of mesenchymal stem cell differentiation via the FTO-MMP1-ERK pathway. Similarly, Xin *et al.* linked decreased ESR1 expression in concave side muscle progenitor cells to greater fatty infiltration and scoliosis severity (29), supporting the adipogenic shift observed in our study. In our dataset, fetal Schwann cell signatures were positively enriched in the scoliosis cases. This contrasts with the morphological reduction of Schwann cells in the scoliosis cases shown by Repko *et al* (*30*), although that study did not reflect the same approach and cell developmental stage. We also observed increased satellite cell gene expression on the concave side, in line with recent finding of higher PIEZO2 satellite cells numbers on the concave side in adolescent idiopathic scoliosis (31) and mechanical stretching being known to activate satellite cells (32). In addition, *CXCL14* was upregulated on the convex side, consistent with its roles in muscle regulation. This chemokine has been shown to be enriched in fetal and juvenile muscle-related cells. Previous research has shown that *CXCL14* negatively regulates skeletal muscle regeneration by preventing myoblasts from exiting the cell cycle, which is necessary for differentiation (33). This might have a stronger effect upon aging and has been linked to reduced muscle repair capacity (34). The altered expression of *CXCL14* in our data may reflect a similar mechanism, where muscle regeneration or differentiation is disrupted, potentially contributing to the observed asymmetry between the two sides of the spine. Finally, genes associated with T cells, B cells, myeloid cells and NK cells were downregulated in scoliosis samples, suggesting reduced immune activity in the paravertebral muscle tissue. This finding supports that immune dysregulation contributes to the asymmetric tissue remodeling observed in scoliosis.

To better understand whether the observed transcriptional asymmetry may originate from developmental processes and to examine the patterns of gene regulation across developmental stages, we analyzed the expression of DEGs across embryonic, fetal, juvenile and adult stages using the UCSC Cell Browser data (12, 13). We observed that *CXCL14, JUNB, LDHB, LUM, PTN, RPL27* are highly expressed during specific fetal stages and in specific cell types. Notably, *JUNB* has been previously identified as part of a key regulon across multiple developing organs between PCW3-PCW8 in the human embryonic organogenesis atlas (Pan et al., 2023, Supplementary Table 3), highlighting its early developmental regulatory role. Similarly, *ACTC1*, a structural muscle gene, was upregulated on the convex side. Although *ACTC1* itself was not specifically discussed in the recent human embryonic organogenesis atlas (35), closely related members of actin family such as *ACTA1*, *ACTB* and *ACTG1* showed specific temporal expression patterns during human muscle and cardiac development at post-conceptional weeks (PCW) 3 to PCW8 (35) (Figure 2K). In addition, findings from Pan *et al.* showed that *LDHB* (lactate dehydrogenase), while it has not previously been associated with scoliosis, emerges and reaches its peak expression during early organ development at PCW3-5 (35)(Figure 2L), especially in the liver. These expression pattern differences show that several DEGs are active during early developmental stages and may be involved in cell fate specification, suggesting an early developmental origin in the observed asymmetry. Similar developmental timing patterns have been reported in previous studies for adolescent idiopathic scoliosis. For example, Burwell *et al.* proposed a Cascade Concept, that scoliosis may originate from disruptions in cell types of different embryonic origins, including adipocytes and osteoblasts derived from mesenchymal stem cells, and myoblasts potentially triggered by environmental or epigenetic factors (25). Similarly, Zaydman *et al.* suggested that altered Pax3-mediated neural crest cell migration may contribute to early musculoskeletal imbalances (36), while Jinnah *et al.* proposed that asynchronous neuro-osseous growth mechanisms originating in early developmental asynchrony may play a central role (37). Moreover, prospective studies show differences in body composition identifiable before the clinical onset of scoliosis, reinforcing the hypothesis that scoliosis-related tissue divergence begins early in life (38).

A major strength of our study is the inclusion of a comparatively large cohort of both scoliosis cases and controls. Among limitations are that samples were obtained from different levels in scoliosis cases and controls; and group differences in mean age and sex distribution. Additionally, scoliosis samples were collected from both primary and compensatory scoliosis curves.

Taken together, our study extends previous observations of paravertebral muscle asymmetry by integrating transcriptomic, radiological and cell type enrichment analyses. We show that the convex side has a more muscle related pattern whereas the concave side has a more adipose related pattern, associated with immune and extracellular matrix changes. Together, these findings suggest that the molecular differences observed in scoliosis may originate during early development.

## Methods

### Sex as a biological variable

In this study, sex was not considered as a biological variable.

### Study participants

The flowchart for inclusion and exclusion is presented in Figure 1. Patients with diagnosed idiopathic scoliosis, aged 10-25 years, undergoing surgery at the Karolinska University Hospital, Sweden between 2018 and 2021 were invited to participate in the study. Inclusion was done consecutively. Patients of similar age undergoing spine surgery at Karolinska University hospital for non-scoliotic conditions participated as controls (Figure 1).

### Radiology

The preoperative Cobb angle was determined using the last erect whole spine radiograph before surgery for the idiopathic scoliosis cases (39). The convexity and the levels of sample collection for each scoliosis case is shown in the Supplementary Figure 4. For the controls, erect spine x-ray was not always performed, however computed tomography (CT) and magnetic resonance images (MRI) were examined to rule out any scoliosis curves.

The preoperative whole spine CT was available for 26 out of 35 scoliosis cases and examined. Measurements of Hounsfield units (HU) on the paravertebral muscle on the level of muscle biopsy was performed as described by Shimoda *et al.* (40). A third-year resident in orthopedic surgery (TC) and an orthopedic spine surgeon with 25 years of experience in spinal deformity surgery (PG) made the measurements independently of each other.

Each observer made multiplanar reconstructions of the computed tomography for a “true axial” of the spine at the level of the muscle biopsies. All measurements were made with the imaging software SECTRA IDS7 version 25.2 (SECTRA, Linköping, Sweden). The mean of the two observers’ measurements for HU for each patient on the convex side and concave side was calculated and presented using a paired dot plot. The Wilcoxon signed-rank test was performed using RStudio version 2023.23.2+402.

### Sampling

Muscle biopsies were obtained from both the concave and convex sides of the scoliosis cases during surgery. The muscle biopsies were obtained predominantly near the convexity of the major curve or in the convexity of the compensatory curve. The level of sampling and the convexity of the curve is shown in Supplementary Figure 4 for all scoliosis cases. For the controls, the majority had spinal conditions requiring surgery in the lumbar spine. Since not all patients underwent surgery on both sides of the spine, a single-sided sample was obtained when bilateral samples could not be collected (Supplementary Figure 4). Muscle biopsies were obtained during surgery and transferred directly into RNA*later*™ solution (Invitrogen, Thermo Fisher Scientific) for stabilization. The samples were initially stored in room temperature for 24 hours and subsequently stored long term in –80°C. RNA extraction and sequencing was performed at the same time for all samples.

### RNA extraction

Total RNA was isolated from the tissue samples using the RNAeasy Lipid Tissue Mini Kit (Cat. No. 74804, Qiagen). RNA quality was assessed as RIN scores, using an Agilent BioAnalyser. Concentration of RNA was measured using Qubit.

### RNA-sequencing with STRT method

A modified 5’– end RNA-seq method, Single-Cell Tagged Reverse Transcription (STRT), with 8-bp unique molecular identifiers (UMIs) was applied (18, 41). 40 ng of total RNA from each sample was used for library preparations. Briefly, RNA samples were placed in a 48-well plate in which a universal primer, template-switching oligos, and a well-specific 6bp barcode sequence were added to each well. The synthesized cDNAs from the samples were then pooled into one library and amplified by single-primer PCR with the universal primer sequence. The resulting amplified library was then sequenced using an Illumina NextSeq500 instrument, High Output (75 cycles, single-end), generating approximately 80 million reads per library (1.6–1.8 million reads per sample). Sequences were processed by STRTprep pipeline (41) for the quality check and further analysis.

### Preprocessing and quality control of the STRT libraries

The STRTN pipeline (https://github.com/gyazgeldi/STRTN, commit e16a6d1)(42) was used to preprocess the raw STRT-seq data. Reads were aligned to the human hg38 reference and annotated using GENCODE v43 basic annotation (wgEncodeGencodeBasicV43), which was downloaded from the UCSC Genome Browser at https://genome.ucsc.edu/) (43). After that, for each multiplexed STRT libraries (48 samples per library), a gene count matrix was obtained along with the following quality control metrics: mapped reads, spike-in reads, spike-in 5’-end rate, mapped rate, mapped/spike-in ratio, and coding 5’-end rate metrics. Based on these metrics, for two libraries, outlier samples, non-template controls (NTCs) and technical duplicates were excluded from both libraries.

### Reduction of technical biases and noise in the gene expression profiles

Genes expressed in >5 samples were retained, and their count matrix and depth files were processed to correct for library-specific biases between the two STRT libraries, using the Negative Binomial Generalized Linear Model – Library Bias Correction (NBGLM-LBC) package in R (version 4.2.2)(44). Depth files, extracted from BAM files using samtools (45), were processed and classified by library. A feature selection test was then used to identify genes with high variability compared to spike-in noise; genes with adjusted p-values <0.05 were considered fluctuating (41). Principal Component Analysis (PCA) was performed on both the raw count matrix and library bias corrected count matrix using the Seurat package (version 5.0.1) in R to identify library bias, followed by spike-in normalization with log transformation (46).

### Differential gene expression (DGE) analysis

DGE was analyzed using the DESeq2 package (version 1.38.3)(47) with Wald test in R. Spike-in normalization was performed by specifying the spike-ins in the controlGenes parameter. The comparisons included 56 scoliosis samples versus 22 control samples, and 30 convex versus 26 concave side samples within the scoliosis group. Library information was used as a covariate factor in the model design. The results were visualized using the EnhancedVolcano package (version 1.16.0)(Blighe, 2018/2024) in R.

### Gene set enrichment analysis (GSEA)

GSEA was performed using curated cluster markers for cell types from the C8 collection, to identify enriched cell types in the sample groups (48–50). The ranking of genes based on log2FC and p-value was analyzed using the fgsea package (version 1.24.0) in R (51). For this analysis, gene sets related to muscle and fetal muscle development were selected from “DESCARTES FETAL MUSCLE” (fetal tissues) (26) and “RUBENSTEIN SKELETAL MUSCLE” (adult skeletal muscle, mononuclear cell populations) (27) datasets. The Developmental Single Cell Atlas of gene RegulaTion and ExpresSion (DESCARTES) dataset profiles a single-cell atlas of human fetal tissues (72-129 days post-conception) across 15 developing organs, including fetal skeletal muscle. The RUBENSTEIN dataset profiles adult skeletal muscle (24-84 age), combining RNA-seq of isolated Type I and Type IIa muscle fibers with single-cell RNA-seq of mononuclear cells such as fibro-adipogenic progenitors, satellite cells, endothelial, and immune cells from vastus lateralis biopsies.

### Visualization

Datasets were downloaded from UCSC Cell Browser – the Skeletal Muscle (12) for embryonic development hindlimb muscle (weeks 5-6, 6-7 and 7-8), fetal development hindlimb muscle (weeks 9, 12-14 and 17-18), juvenile hindlimb muscle, and adult hindlimb muscle (13) to explore whether the expression of the significantly changed genes changes during muscle development. The count matrices and associated metadata were combined, and the NormalizeData function from the Seurat package was applied with default parameters to normalize this combined dataset. Following that, a heatmap and violin plot were created to illustrate the expression of differentially expressed genes in the relevant cell populations across developmental stages using ComplexHeatmap package (version 2.14.0) (52) and the VlnPlot function of Seurat. UMAPs from the UCSC-Cell Browser website were also used to show the expression profiles across these developmental stages (12).

### Study approval

This study was approved by the Swedish Ethical Review Authority in Stockholm. Case (Dnr) number 2017/2374-31, 2018/1457-32, 2022/00700-02, 2024/05452-02. Written informed consent were obtained from all study participants prior to participation in the study.

### Data availability

The datasets generated in this study include sensitive information, therefore are not publicly available due to the agreement of informed consent. However, access to the data can be granted upon reasonable request to the corresponding author if approved by the ethics committee and the Uppsala University GDPR officer.

## Supporting information

All supplemental files

## Acknowledgements

The authors would like to thank all the patients that participated in the study. We would like to thank Kourosh Jalalpour and Anastasios Charalampidis for help with the biopsies during surgery, and research nurses Luigi Belcastro and Maria Wikzén for assisting with patient inclusion and sample collection. We also appreciate the help from Tiina Skoog, Outi Elomaa and Auli Saarinen for helping us with the sample preparation and transport, MTA and RNA extraction.

The pre-processing analysis was conducted using the ePouta high performance computing platform of CSC (Center for Science, Finland).

