## Supplementary material for "Fetal pathogenesis of scoliosis suggested by asymmetry of gene expression in paravertebral muscles": All supplemental files

### Supplementary Files

**Supplementary Figure 1. Outlier samples were excluded from further analysis based on quality control metrics across both libraries.** (A) In the first library, outliers were samples 6, 15, 33 (NTC), 37, 46, 47, as well as sample 2 due to technical issues and sample 16 as a technical duplicate. (B) In the second library, outliers were samples 3, 5, 7, 8, 9 (NTC), 18, 44, 45, 48, and sample 43 as a technical duplicate.

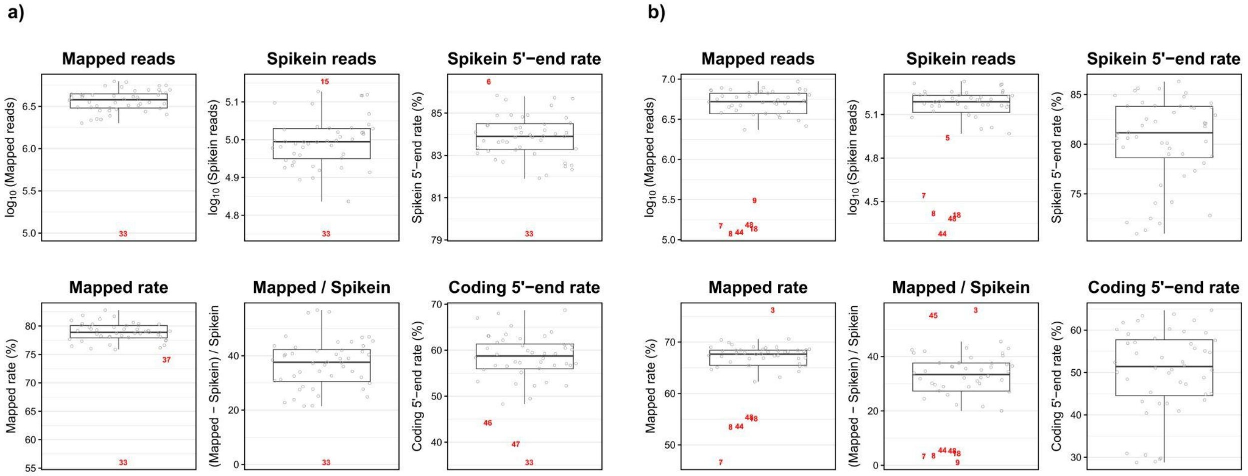

**Supplementary Figure 2. Bias between the two libraries.** (a) Bias before library correction. (b) Existing bias remaining after correction. Due to this, library information as a covariate factor was included in the DGE analysis model.

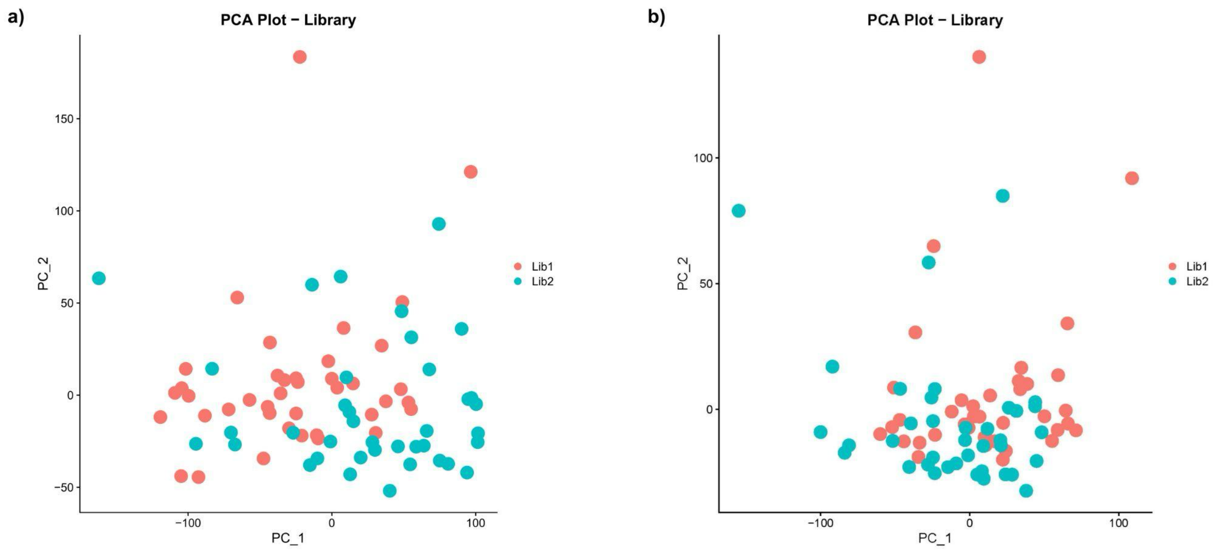

**Supplementary Figure 3. Gene expression dynamics of differentially expressed genes (DEGs across developmental stages and cell types.** Heatmap showing the normalized expression levels (log1p values) of DEGs during embryonic stages in the UCSC-Cell Browser Skeletal Muscle Datasets. Columns represent the developmental stages: embryonic, fetal, juvenile, and adult. Rows correspond to DEGs identified in our analysis. The color scale indicates log1p normalized expression levels, with yellow representing higher expression and purple indicating lower expression. Columns are split by four developmental stages: embryonic, fetal, juvenile, and adult and represent cell types including Chondro (Chondrocytes), PreChondro (Pre-Chondrocytes), Limb.Mesen (Limb Mesenchymal Cells), MSC (Mesenchymal Stem Cells), FAP, FAP.1, FAP.2 (Fibro-Adipogenic Progenitors), Hema (Hematopoietic Cells), EC-Hema (Endothelial-Hematopoietic Cells), RBC (Red Blood Cells), WBC (White Blood Cells), Skin, Dermal, SkM (Skeletal Muscle Cells), SMC (Smooth Muscle Cells), Schwann, EC (Endothelial), and Teno (Tenocytes).

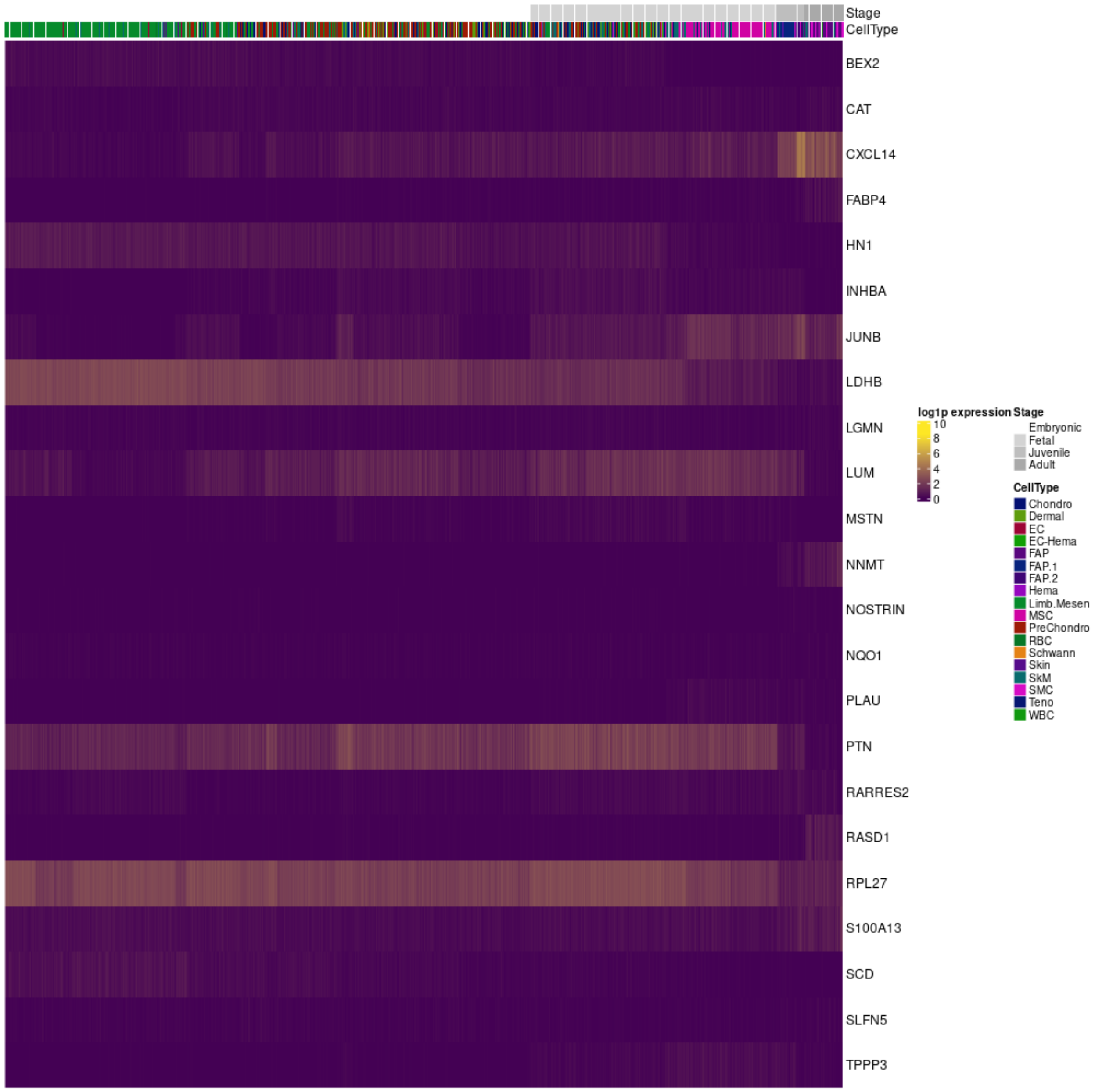
Stage
CellType
BEX2
CAT
CXCL14
FABP4
HN1
INHBA
JUNB
LDHB
LGMN
LUM
MSTN
NNMT
NOSTRIN
NQO1
PLAU
PTN
RARRES2
RASD1
RPL27
S100A13
SCD
SLFN5
TPPP3
log1p expression
10
8
6
4
2
0
Stage
Embryonic
Fetal
Juvenile
Adult
CellType
Chondro
Dermal
EC
EC-Hema
FAP
FAP.1
FAP.2
Hema
Limb.Mesen
MSC
PreChondro
RBC
Schwann
Skin
SkM
SMC
Teno
WBC

**Supplementary Figure 4.** Radiographs of all patients and samples that have been successfully analyzed. The level marking indicates the level where the muscle biopsies were obtained. The triangle indicates what we determined as the convex side while the circle indicates the concave side. Samples that were not analyzed were not shown. For controls, a generic X-ray image of the lumbar spine was used to illustrate the level and side of the sample.

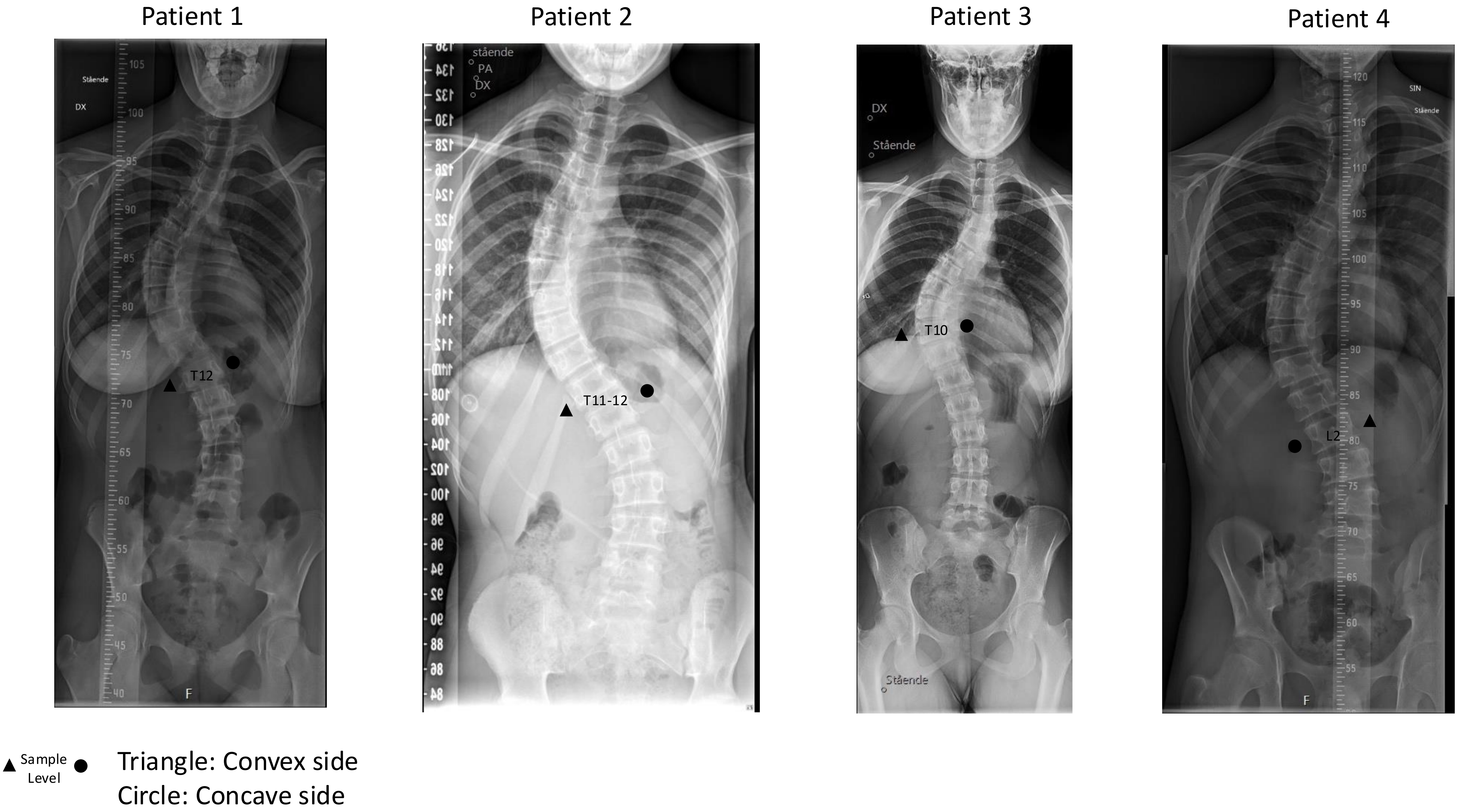

Patient 1
Patient 2
Patient 3
Patient 4
T12
T11-12
T10
L2
Sample Level
Triangle: Convex side
Circle: Concave side

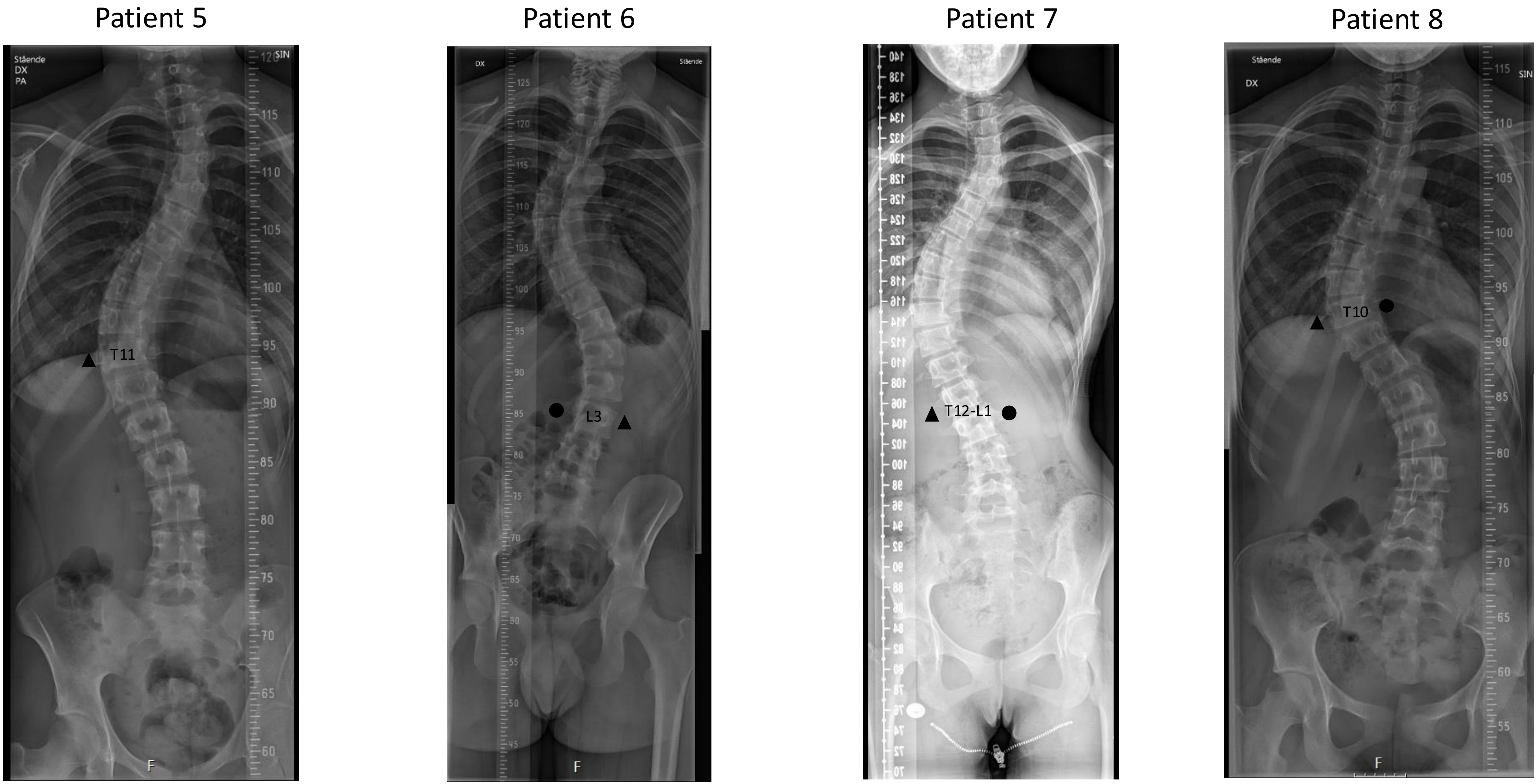
Patient 5
T11
Patient 6
L3
Patient 7
T12-L1
Patient 8
T10

Patient 9

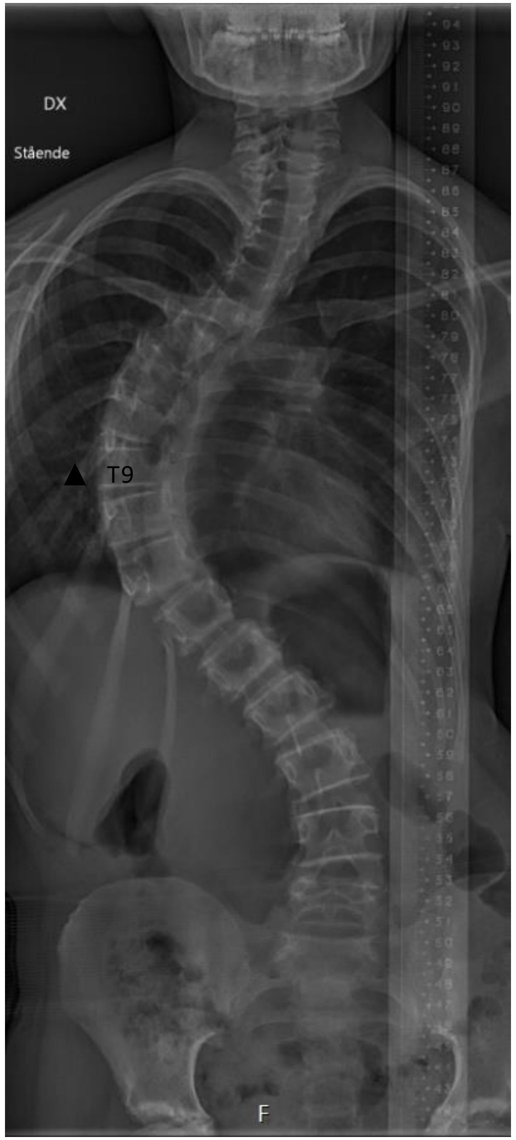


Patient 10

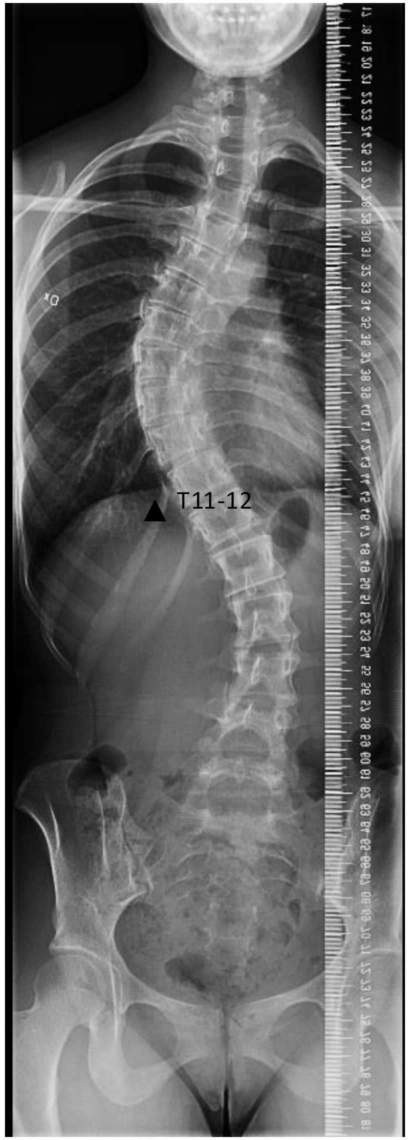


Patient 11

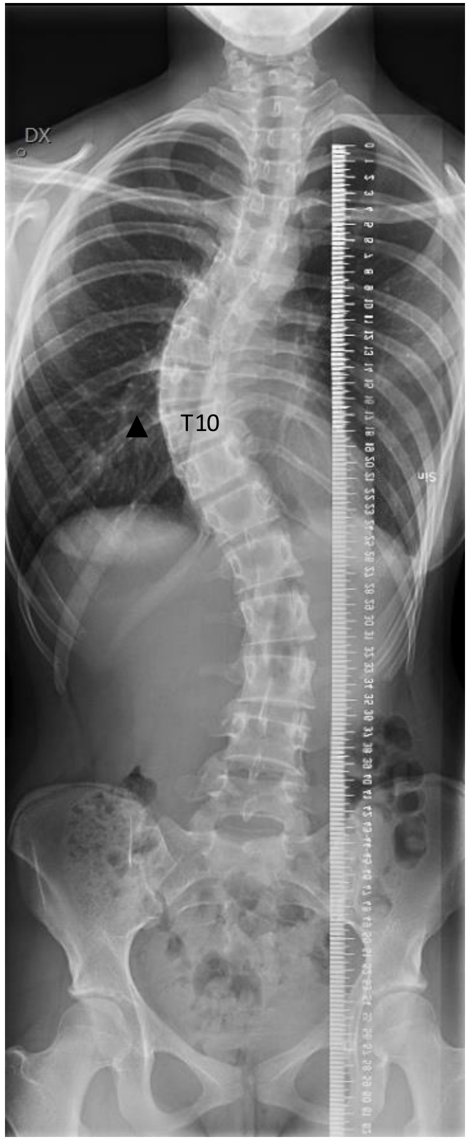


Patient 12

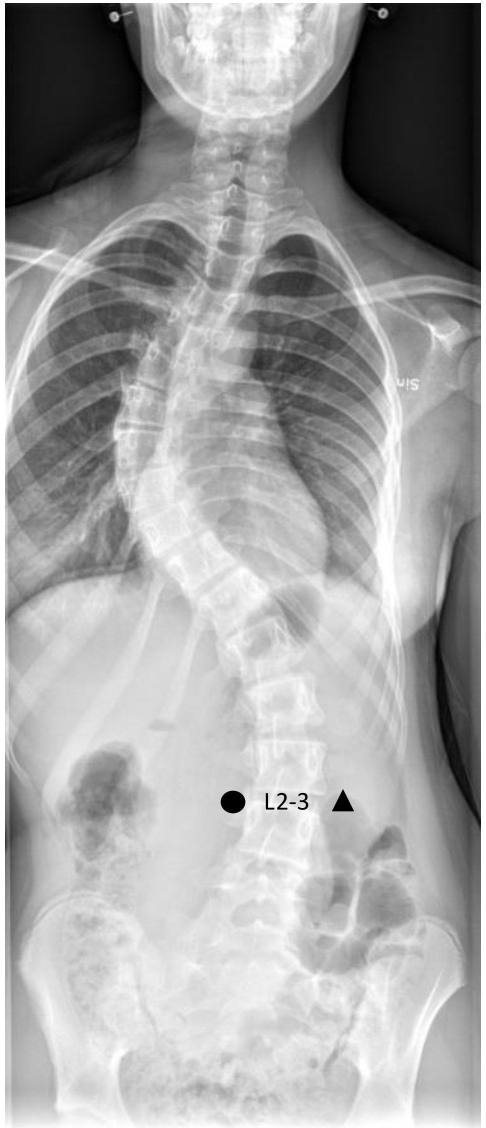

Patient 13

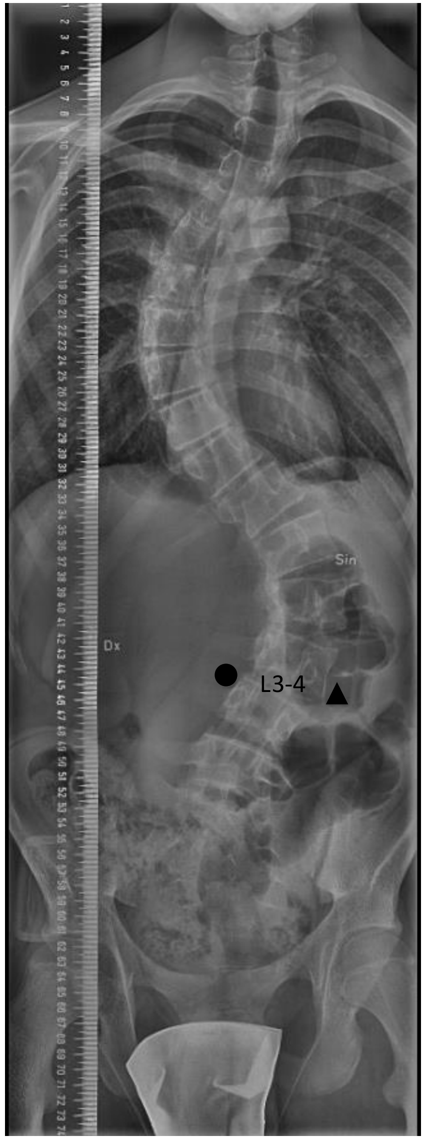


Patient 14

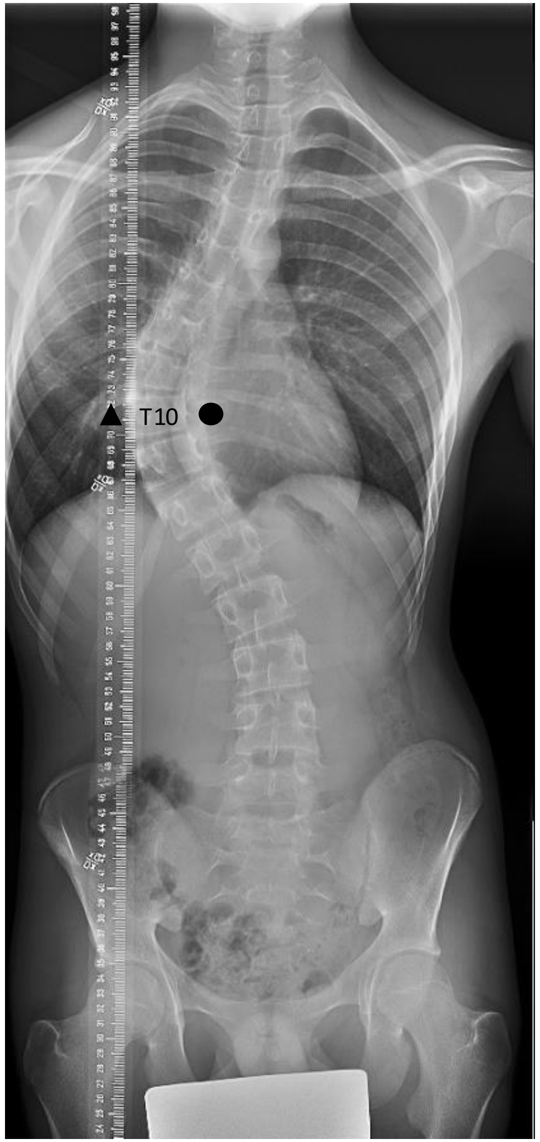


Patient 15

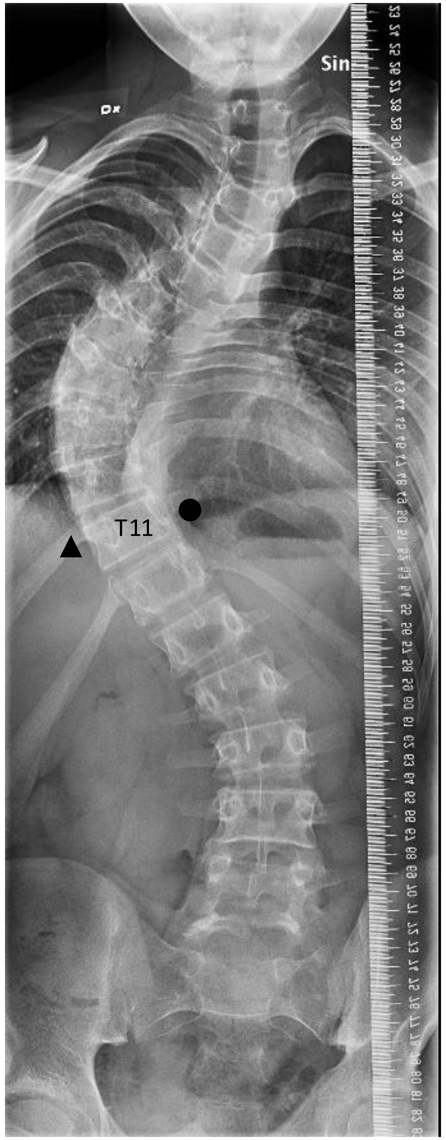


Patient 16

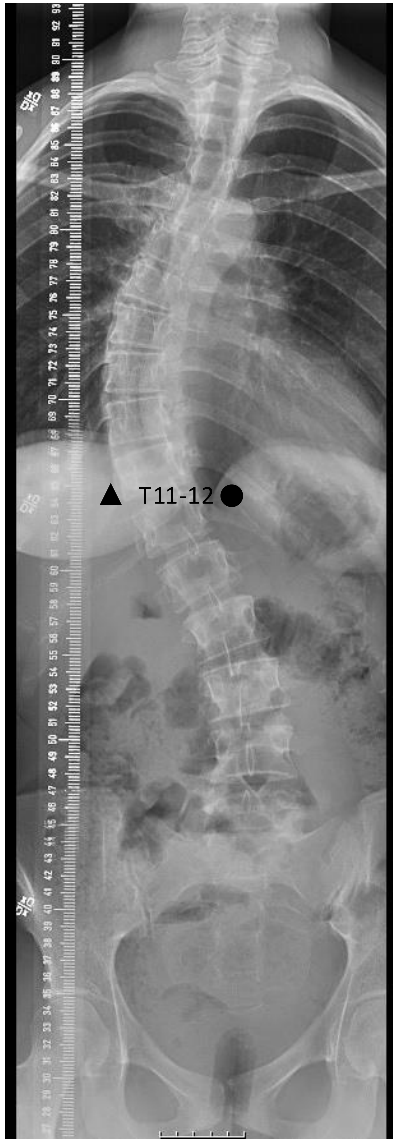

Patient 17

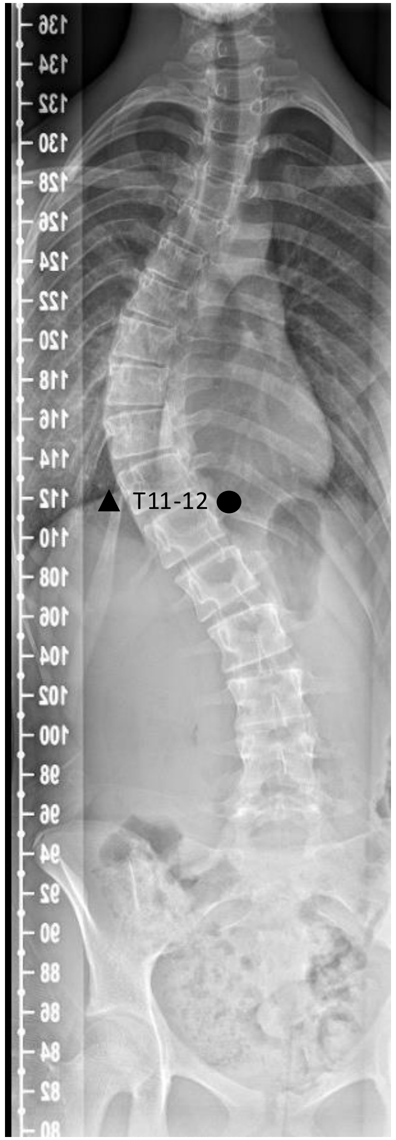


Patient 18

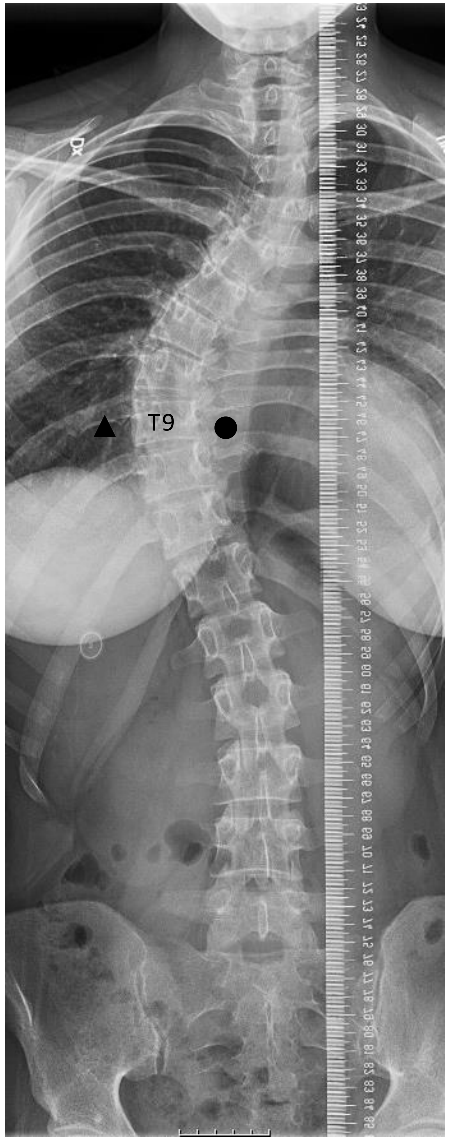


Patient 19

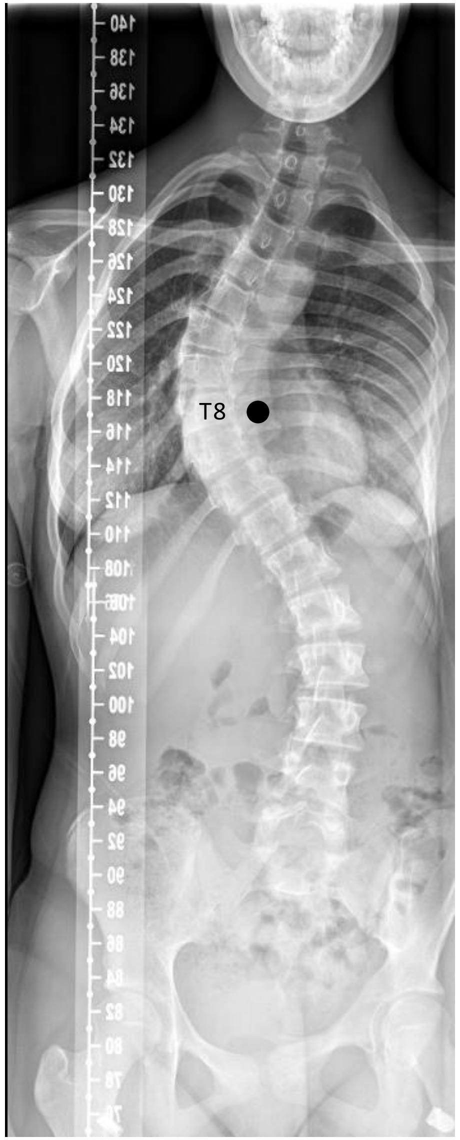


Patient 20

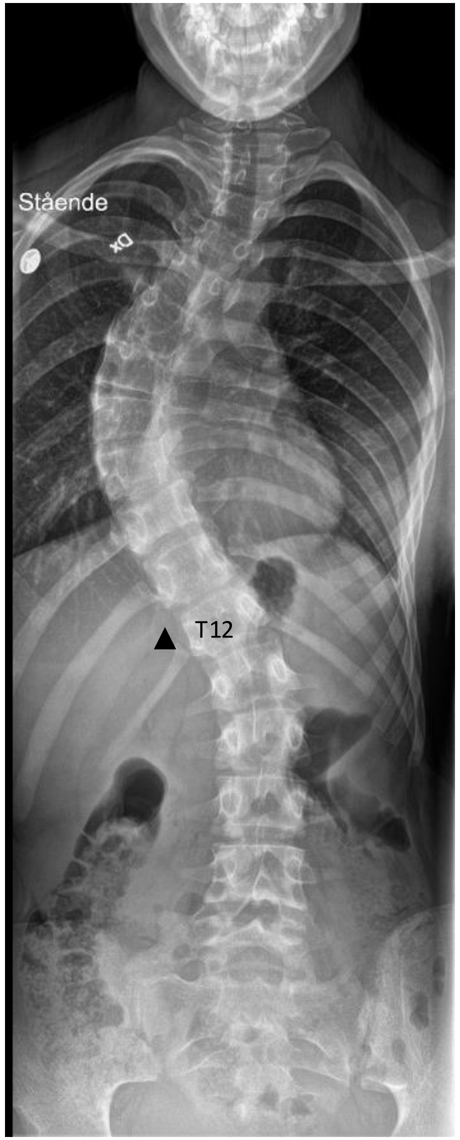

Patient 21

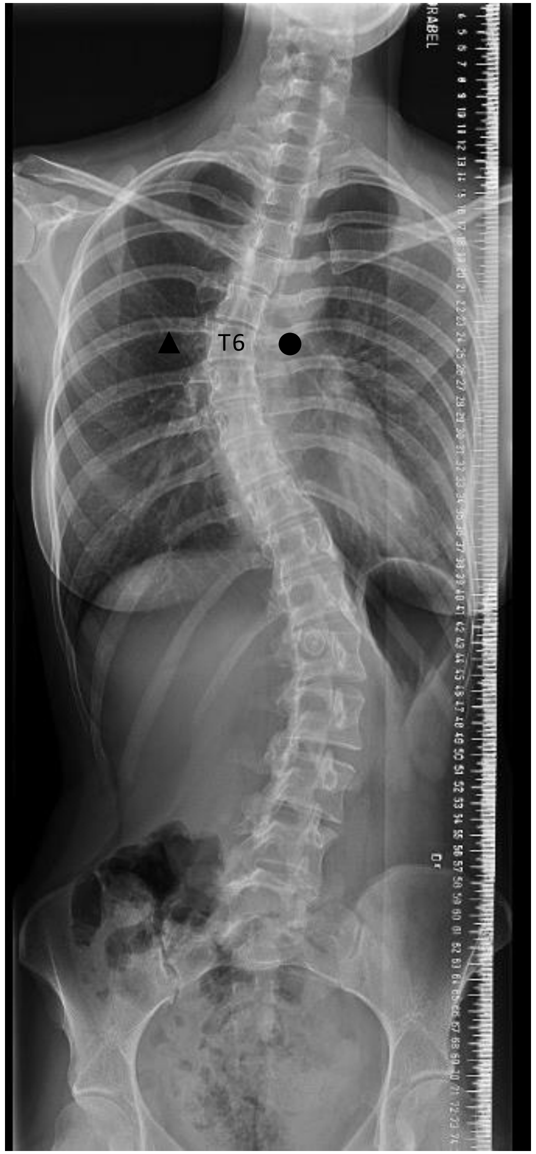


Patient 22

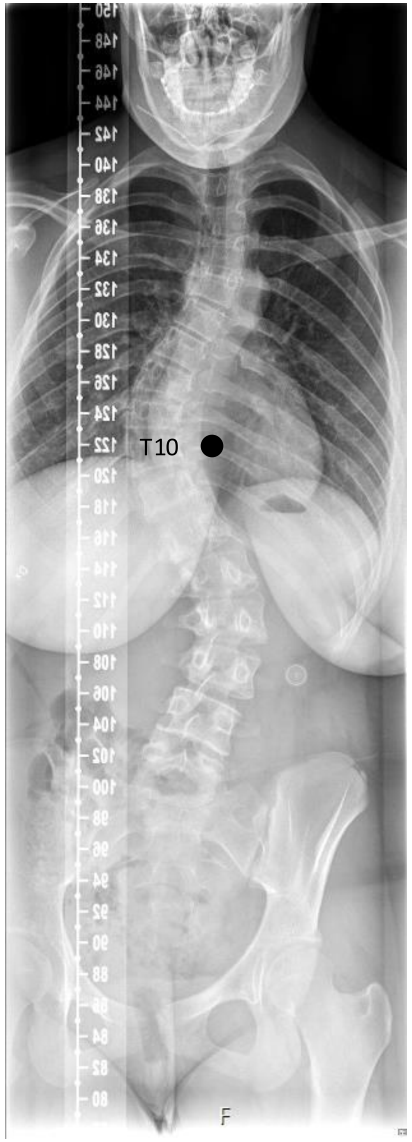


Patient 23

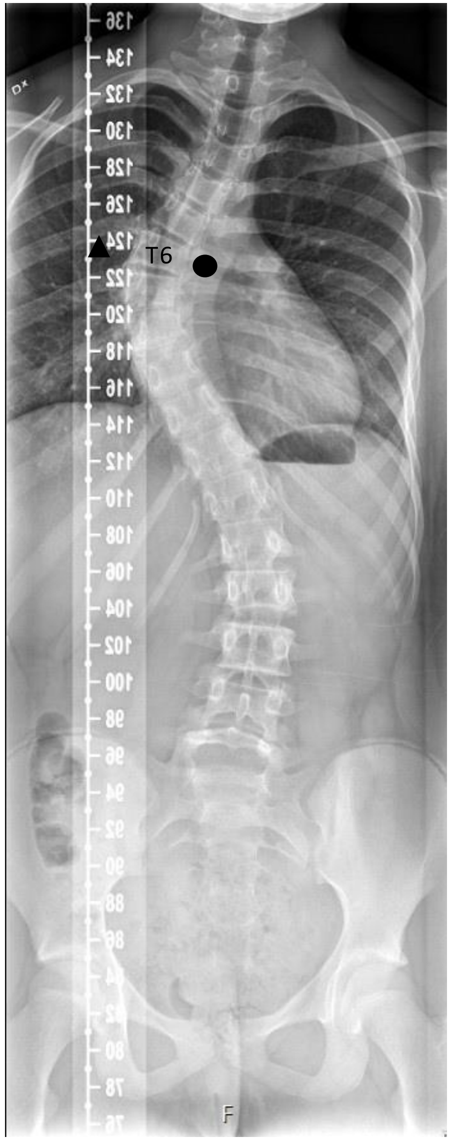


Patient 24

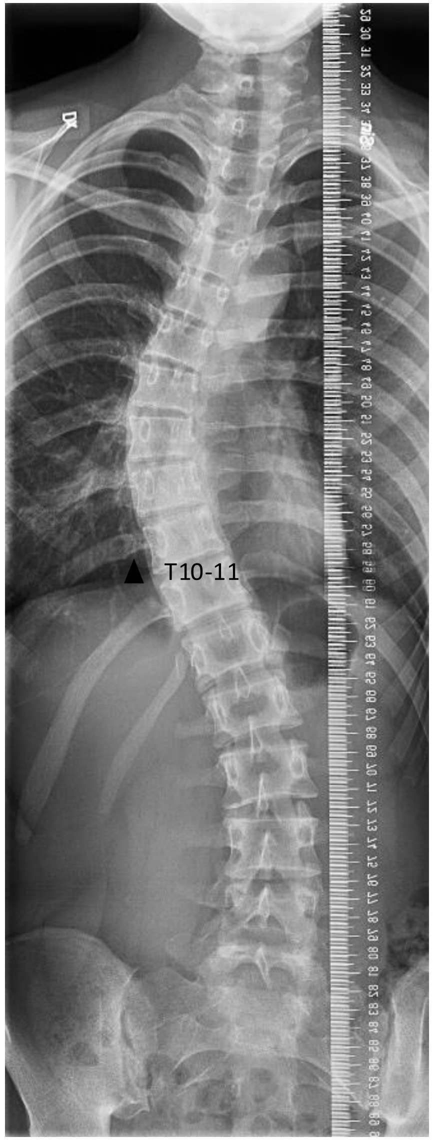

Patient 25

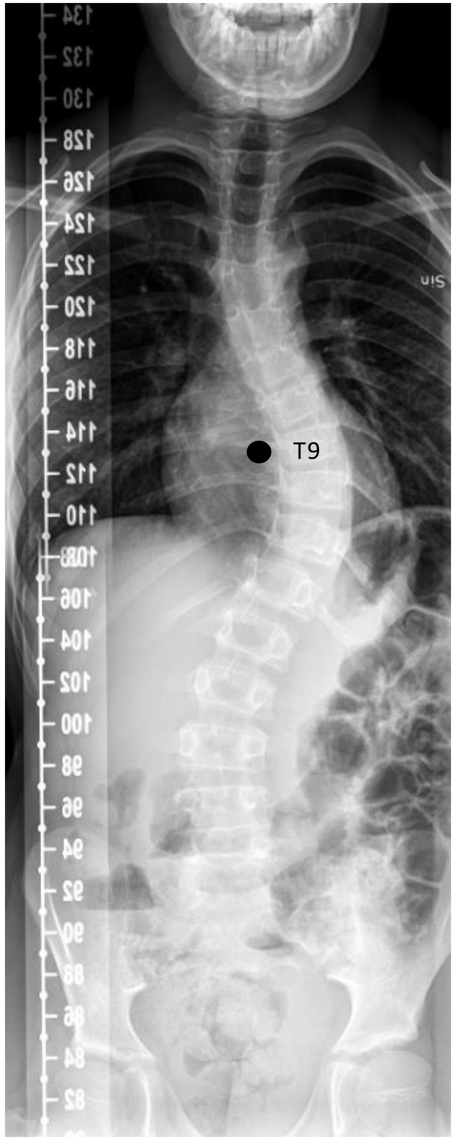


Patient 26

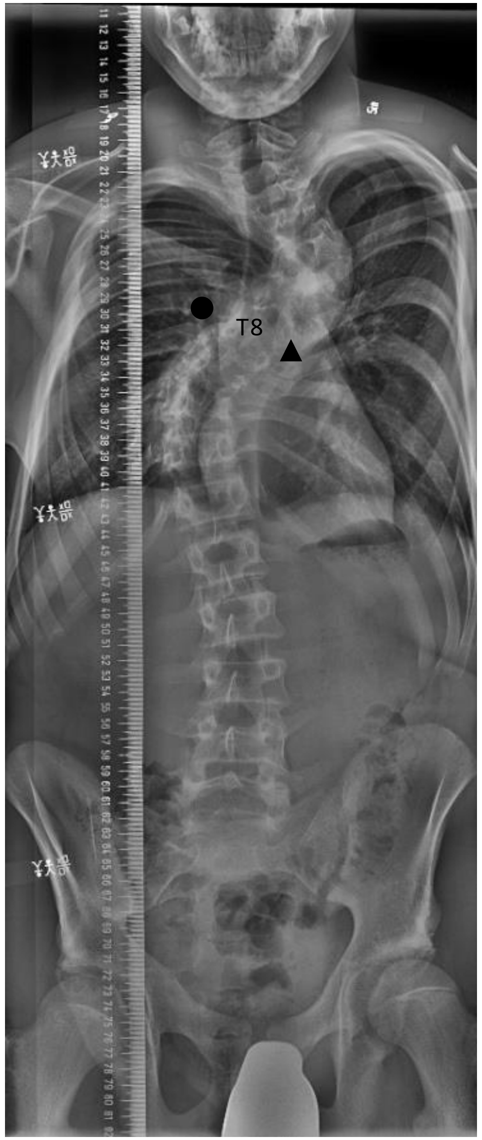


Patient 27

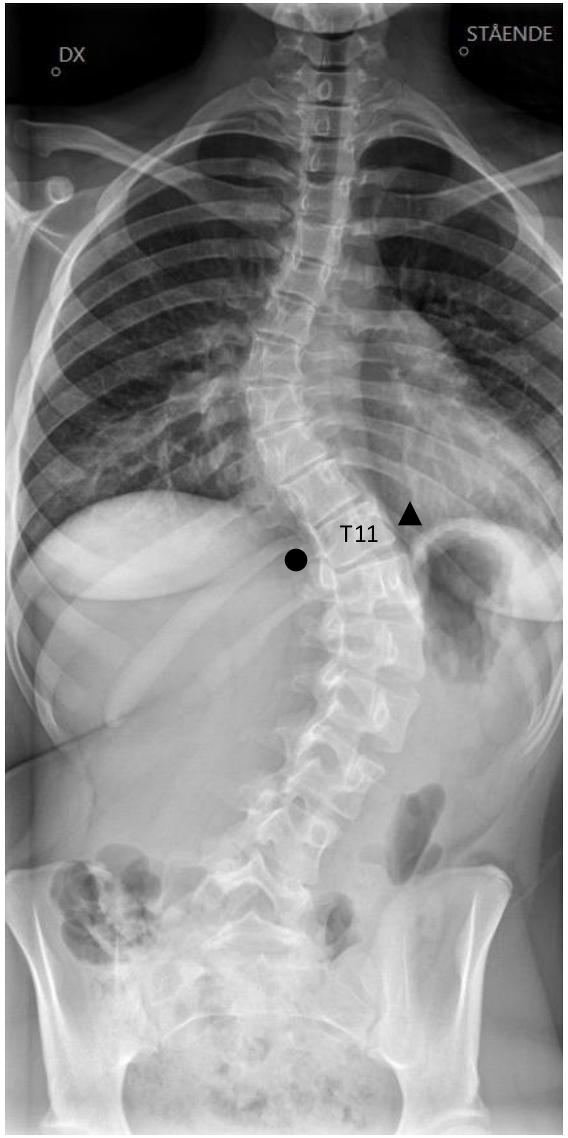


Patient 28

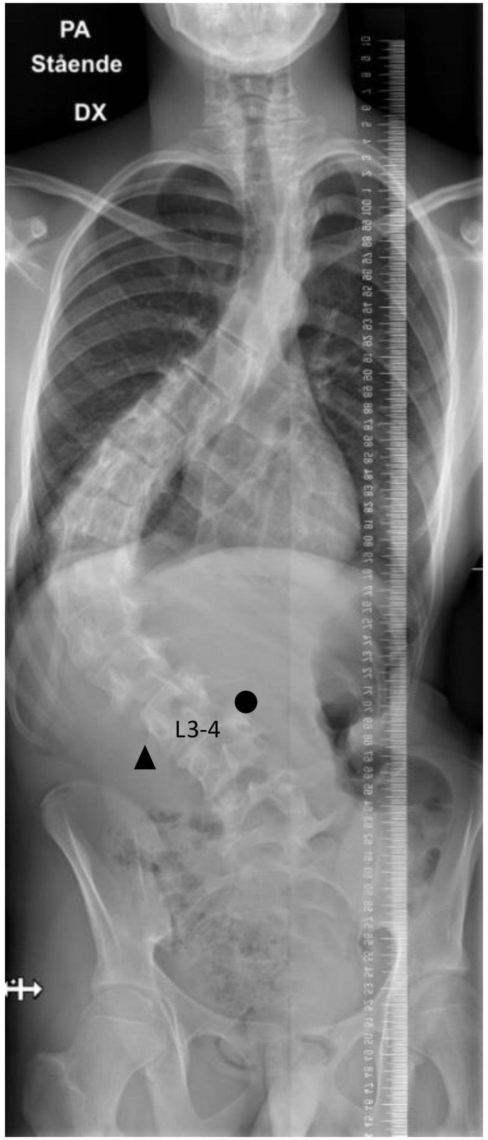

Patient 29

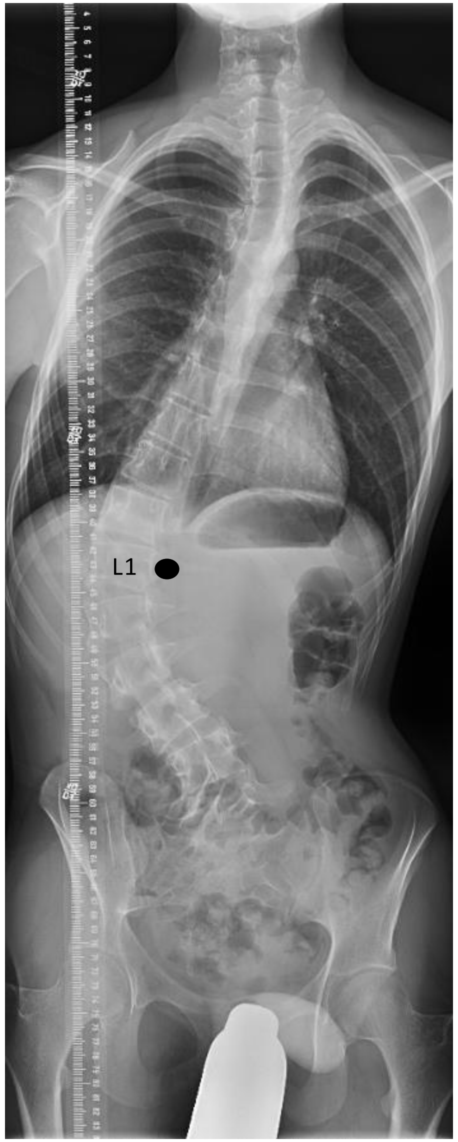


Patient 30

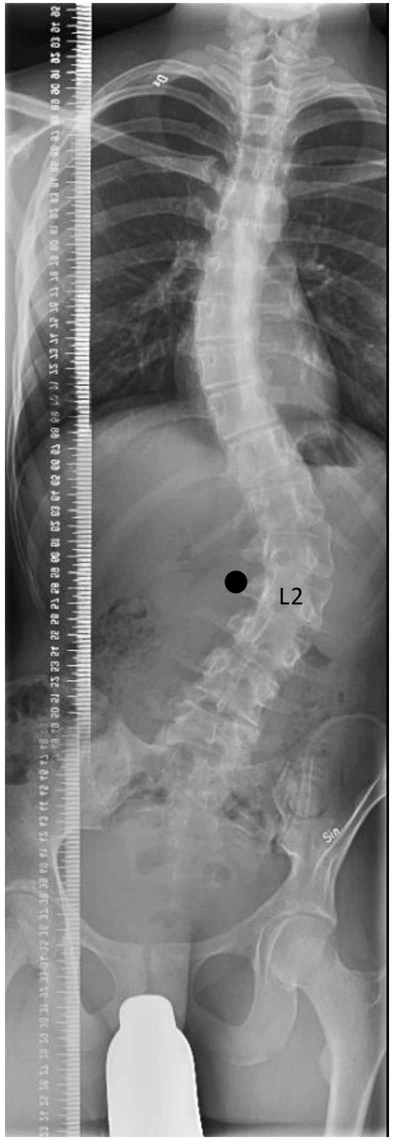


Patient 31

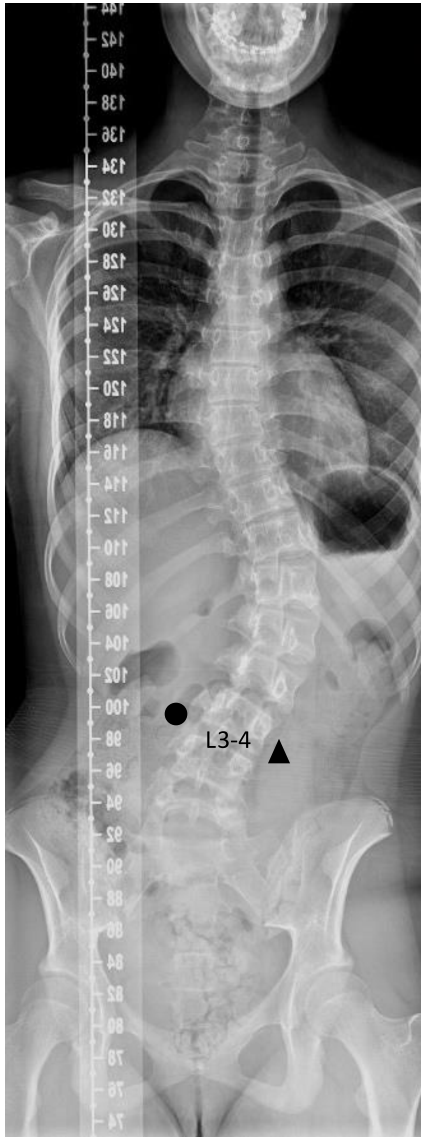


Patient 32

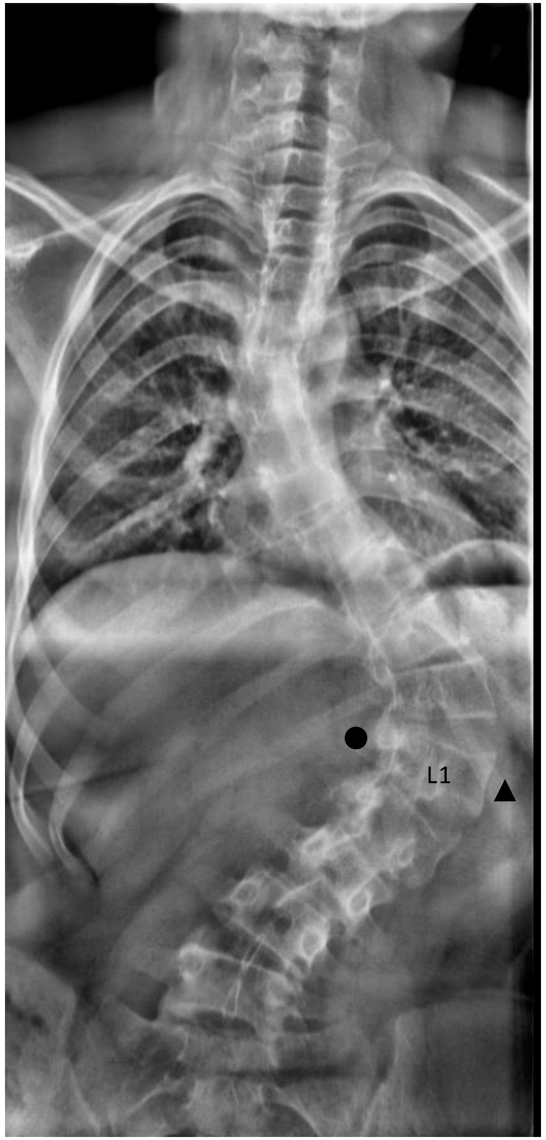

Patient 33

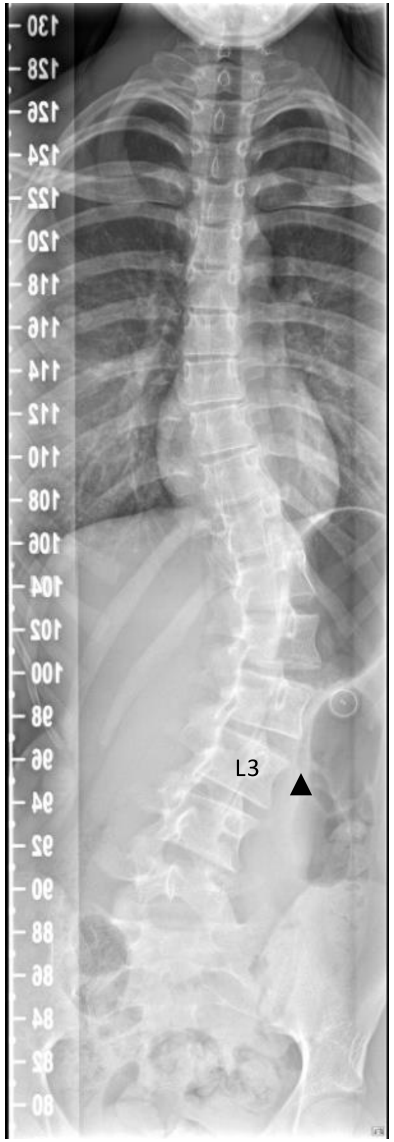


Patient 34

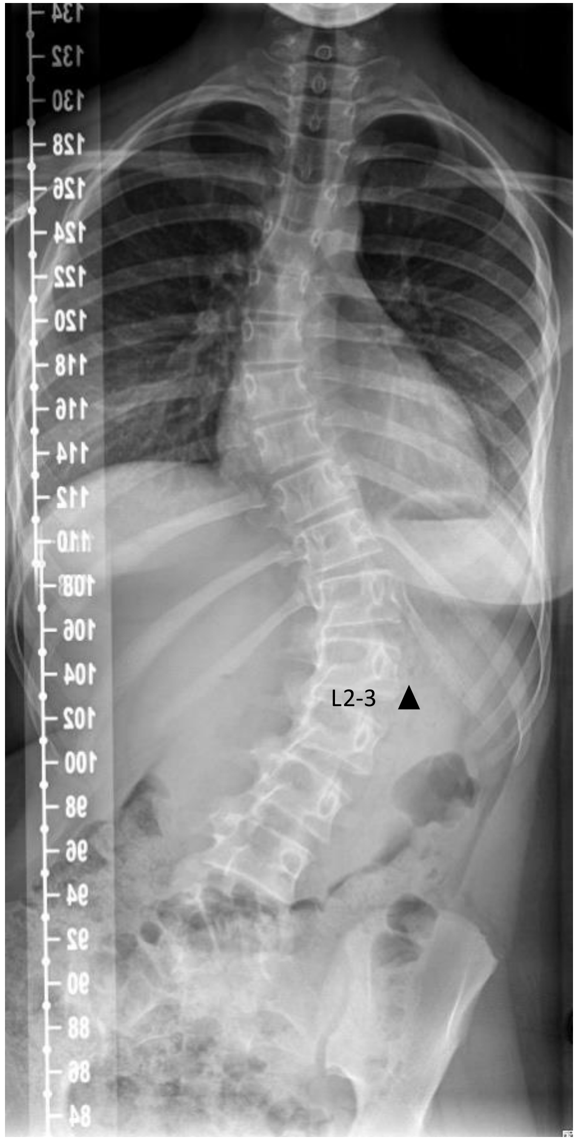


Patient 35

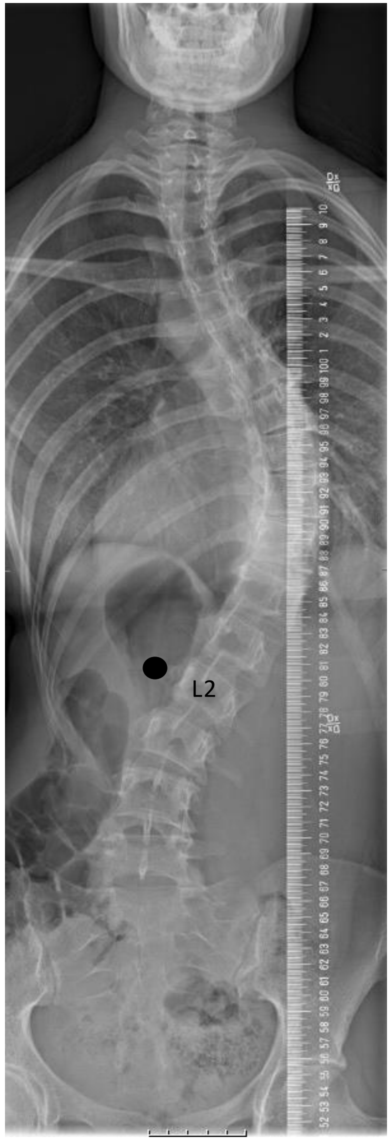

Control bilateral samples n=5

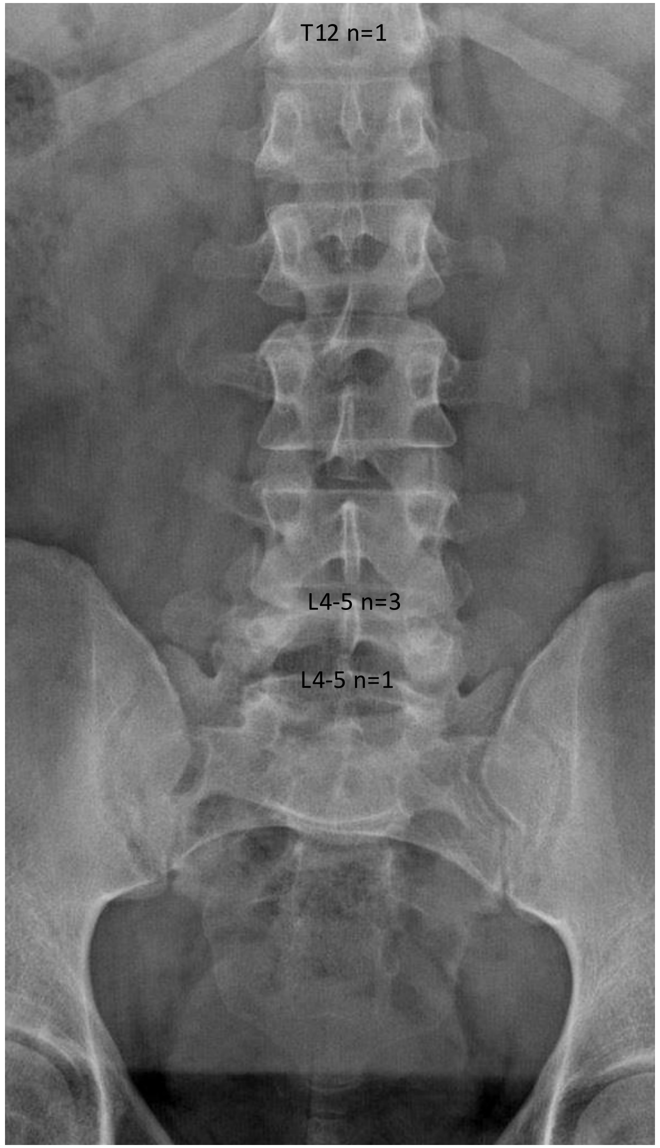


Control left side n=3

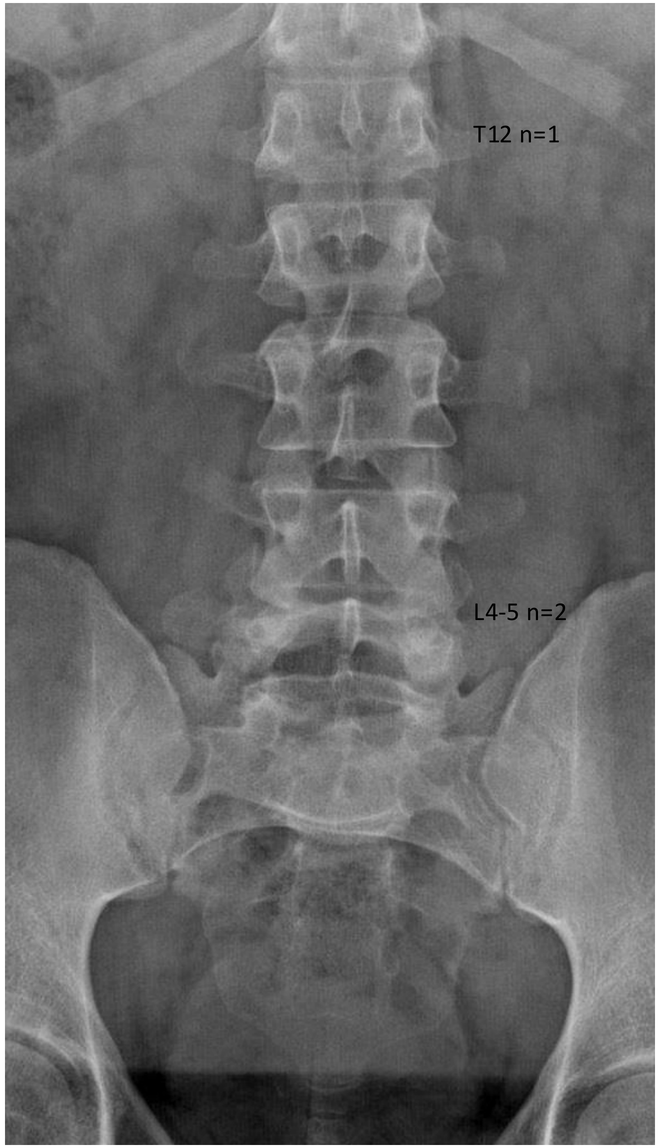


Control right side n=9

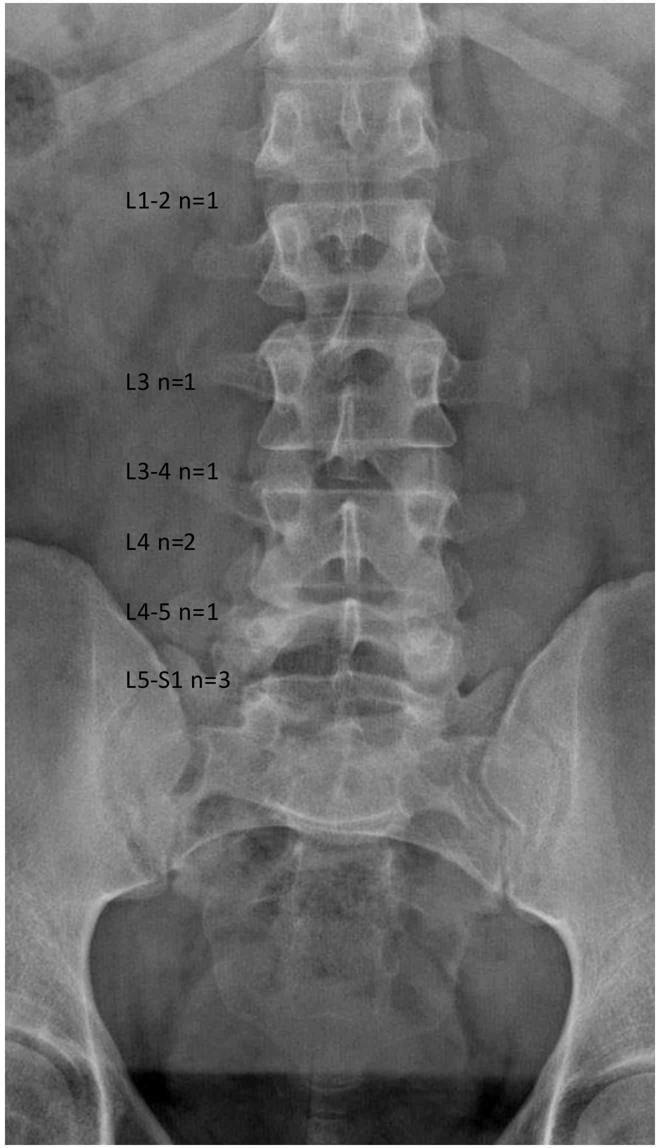

**Supplementary Table 1** – Baseline data for idiopathic scoliosis cases and controls. Data is presented as number (percentage) and mean (standard deviation). n/a: not available

| | Scoliosis Cases (n=35) | Controls (n=17) |
|---|---|---|
| **Sex n (%)** | | |
| Females | 24 (69) | 5 (29) |
| Males | 11 (31) | 12 (70) |
| **Age at surgery m (SD)** | 15.9 (2.4) | 17.1 (2.2) |
| **Diagnosis (n)** | | |
| Idiopathic scoliosis | 35 | 0 |
| Lumbar Disc herniation | 0 | 11 |
| Spondylolisthesis | 0 | 2 |
| Tumor | 0 | 3 |
| Scheuermann's kyphosis | 0 | 1 |
| **Major curve cobb angle m (SD)** | 59 (10) | n/a |
| **Major curve type n (%)** | | n/a |
| Thoracic | 25 (71) | |
| Thoracolumbar | 5 (14) | |
| Lumbar | 5 (14) | |
| **Major curve convexity n (%)** | | n/a |
| Right | 24 (69) | |
| Left | 11 (31) | |
| **Sampling level n (%)** | | |
| T5/T6 & T6 | 2 (5.7) | 0 (0) |
| T7/T8 & T8 | 2 (5.7) | 0 (0) |
| T8/T9 & T9 | 3 (8.6) | 0 (0) |
| T9/T10 & T10 | 5 (14.3) | 0 (0) |
| T10/T11 & T11 | 4 (11.4) | 0 (0) |
| T11/T12 & T12 | 6 (17.1) | 2 (11.8) |
| T12/L1 & L1 | 3 (8.6) | 0 (0) |
| L1/L2 & L2 | 3 (8.6) | 1 (5.9) |
| L2/L3 & L3 | 4 (11.4) | 1 (17.6) |
| L3/L4 & L4 | 3 (8.6) | 3 (41.2) |
| L4/L5 & L5 | 0 (0) | 7 (17.6) |
| L5/S1 & S1 | 0 (0) | 3 (17.6) |

**Supplementary Table 2. Differentially expressed genes in scoliosis muscle samples compared to controls and between convex and concave sides.** The list includes down and up-regulated genes with their descriptions and functions.

| Gene name | Description | Function | Direction of gene expression |
|---|---|---|---|
| *ANGPTL7* | Angiopoietin-related protein 7 | Promotes angiogenesis and extracellular matrix remodeling (1). | Scoliosis > Control |
| *NNMT* | Nicotinamide N-methyltransferase | Catalyzes nicotinamide methylation and regulates NAD+ metabolism and cellular methylation potential. *NNMT* is highly expressed in metabolically active tissues and skeletal muscles. It modulates inflammation, oxidative stress responses, and myogenesis (2, 3). | Scoliosis > Control |
| *INHBA* | Inhibin subunit A | Member of the TGF-β superfamily. | Scoliosis > Control |
| *CNN1* | Calponin 1 | Actin filament binding protein, regulates smooth muscle contraction and proliferation. | Scoliosis > Control |
| *PTN* | Pleiotrophin | Heparin-binding growth factor, involved in cell proliferation, angiogenesis, and tissue remodeling. | Scoliosis > Control |
| *COMP* | Cartilage oligomeric matrix protein | Extracellular matrix protein important for skeletal tissue integrity and cell–matrix interactions. | Scoliosis > Control |

| | | | |
|---|---|---|---|
| *TF* | Transferrin | Iron-binding glycoprotein regulating iron homeostasis and oxidative stress. | Scoliosis > Control |
| *RASD1* | Ras related dexamethasone induced 1 | Small GTPase induced by dexamethasone, linked to cell morphology, growth and cell-extracellular matrix interactions. | Scoliosis > Control |
| *JUNB* | JunB proto-oncogene | Transcription factor, part of AP-1 complex; regulates gene expression in response to growth factors and immune signals. | Scoliosis > Control |
| *HBG1* | Hemoglobin subunit gamma-1 | Part of fetal hemoglobin, helps binding oxygen and iron ion. | Scoliosis < Control |
| *S100A13* | S100 calcium binding protein A13 | Involved in protein export and stress response; plays a role in signal transduction and cell regulation. | Scoliosis < Control |
| *PLAU* | Plasminogen activator urokinase | Serine protease that activates plasminogen to plasmin, involved in tissue remodeling, fibrinolysis, and immune responses. | Scoliosis < Control |
| *JPT1* | Jupiter microtubule associated homolog 1 | Microtubule-associated protein, helps organize the cell skeleton. | Scoliosis < Control |
| *LUM* | Lumican | Extracellular matrix protein, important for collagen structure. | Scoliosis < Control |
| *RPL27* | Ribosomal protein L27 | Catalyzes protein synthesis, needed for proper rRNA processing and maturation of rRNAs. | Scoliosis < Control |

| | | | |
|---|---|---|---|
| *C2* | Complement c2 | Part of C3 and C5 convertase, important for immune defense and pathogen clearance. | Scoliosis < Control |
| *SLFN5* | Schlafen family member 5 | Involved in ATP binding activity and hematopoietic cell differentiation. | Scoliosis < Control |
| *LGMN* | Legumain | Lysosomal cysteine protease, processes antigens for MHC class II presentation and involved in protein degradation. | Scoliosis < Control |
| *IDO1* | Indoleamine 2,3-dioxygenase | Immunomodulatory enzyme that suppresses T and NK cells, promotes immune tolerance, and contributes to angiogenesis in cancer (4). Zhang et al. (5) reported differential *IDO1* expression in lung tissue in an early-onset scoliosis and thoracic insufficiency syndrome model. | Scoliosis < Control |
| *NQO1* | NAD(P)H quinone dehydrogenase 1 | Detox enzyme, protects cells from oxidative stress and supports antioxidant defense. | Scoliosis < Control |
| *TMEM140* | Transmembrane protein 140 | Membrane protein, exact function not well known. | Scoliosis < Control |
| *SYCE1* | Synaptonemal complex central element protein 1 | Involved in meiosis and helps chromosomes pair correctly. | Scoliosis < Control |

| | | | |
|---|---|---|---|
| *GLYAT* | Glycine-N-acyltransferase | A mitochondrial enzyme that plays a vital role in cellular detoxification by catalyzing the conjugation of glycine to toxic metabolic acids, facilitating their solubility and excretion. It has been previously associated with bone size and lean muscle mass in GWAS study, suggesting its potential role in musculoskeletal development (6). | Convex < Concave |
| *THRSP* | Thyroid hormone responsive | Regulates *de novo* lipogenesis and lipid metabolism; associated with fatty acid synthesis, adipogenesis, and energy balance in metabolically active tissues. It has been previously linked to lipogenic gene regulation and muscle mitochondrial gene expression, supporting its role in adipose-muscle metabolic crosstalk (7, 8). | Convex < Concave |
| *CXCL14* | C-X-C motif chemokine ligand 14 | Involved in immune cell recruitment and negative regulation of skeletal muscle differentiation (9). | Convex < Concave |
| *S100B* | S100 calcium binding protein B | Involved in muscle stress response and regeneration. | Convex < Concave |
| *MSTN* | Myostatin | Negative regulator of skeletal muscle growth, inhibits muscle cell proliferation and differentiation. | Convex < Concave |

| | | | |
|---|---|---|---|
| *SCD* | Stearoyl-CoA desaturase | Enzyme for fatty acid metabolism and energy balance. | Convex < Concave |
| *FABP4* | Fatty acid binding protein 4 | Binds fatty acids, important in fat metabolism and adipocyte function (10). | Convex < Concave |
| *NOSTRIN* | Nitric oxide synthase trafficking | Regulates nitric oxide synthase and blood vessel function. | Convex < Concave |
| *RARRES2* | Retinoic acid receptor responder 2, chemerin | Adipokine, involved in immune signaling and fat cell differentiation. | Convex < Concave |
| *ENSG00000173366* | Novel Twinfilin | Actin-binding protein, function not well defined. | Convex > Concave |
| *SCGB1D2* | Secretoglobin family 1D member 2 | Small secreted protein from the lipophilin family regulated by steroid hormones, may bind androgens and estrogens. | Convex > Concave |
| *BEX2* | Brain expressed X-linked 2 | Regulates cell cycle and apoptosis; role in tumor suppression and cancer cell survival. | Convex > Concave |
| *ACTC1* | Actin alpha cardiac muscle 1 | Major actin isoform in cardiac and skeletal muscle, important for contraction. | Convex > Concave |
| *TPPP3* | Tubulin polymerization | Regulates microtubule dynamics and cell shape, important for cell structure and implantation. | Convex > Concave |

| | | | |
|---|---|---|---|
| | promoting protein family member 3 | | |
| *IL18* | Interleukin 18 | Pro-inflammatory cytokine, activates immune cells like NK and T cells. | Convex > Concave |
| *LDHB* | Lactate dehydrogenase B | Enzyme in energy metabolism, converts lactate to pyruvate. | Convex > Concave |

Supplementary Table 2 References:
